# In Their Own Words: Noise Complaint Data Reveals Impacts of Military Aviation

**DOI:** 10.64898/2026.04.14.26350904

**Authors:** Ching-Hsuan Shirley Huang, Lauren M. Kuehne, Giordano Jacuzzi, Julian D. Olden, Edmund Seto

## Abstract

Military aviation training noise remains understudied despite its widespread impacts across urban, rural, and wilderness areas. The predominance of low-frequency noise and repetitive training can create pervasive noise pollution, yet past research often fails to capture the full range of health and quality-of-life effects. This study analyzed two complaint datasets related to Whidbey Island Naval Air Station noise: U.S. Navy records (2017–2020) and Quiet Skies Over San Juan County data (2021–2023). We analyzed and mapped sentiment intensity from noise complaints relative to modeled annual noise exposure, developed a typology to classify impacts, and modeled the environmental and operational factors influencing complaints.

Findings revealed widespread negative sentiment and anger, often beyond the bounds of estimated noise contours, suggesting that annual cumulative noise models inadequately estimate community impacts. Complaints consistently highlighted sleep disturbance, hearing and health concerns, and compromised home environments due to shaking, vibration, and disruption of daily life. Residents also reported significant social, recreational, and work disruptions, along with feelings of fear, helplessness, and concern for children’s well-being. The number of complaints were strongly associated with training schedules, with late-night sessions being the strongest predictor. A delayed response pattern suggests residents reach a frustration threshold before filing complaints.

Overall, our findings demonstrate persistent negative sentiment and diverse impacts from military aviation noise. Results highlight the need for improved noise metrics, modeling and operational adjustments to mitigate the most disruptive effects.

## 1. Introduction

The rise and continued expansion of military aviation since World War II has prompted a growing need to address the human health and well-being consequences of noise exposure from aircraft training activities. The public health challenges of populations residing in proximity to the noise of high-powered jet engines have been documented since the 1950s (Bell, 1956; Borsky, 1961a, 1961b). In fact, early studies of military aviation noise laid the foundation for the assessment and mitigation of commercial (i.e., civilian) aviation noise, following its transition to jet engines (Bell, 1956; Beranek, 1952; Wemheuer, 1979). These early studies identified wide-ranging impacts to health and quality of life as a result of unpredictable and intermittent noise regimes from training activities, including annoyance, disrupted sleep, fear responses, inability to relax or recreate, and increased anxiety and depression (Borsky, 1961a, 1961b). Since then, noise exposure from military aircraft has only expanded in scope and increased in intensity due to higher power engines and afterburners and increased military installations worldwide (Waitz et al., 2005).

Despite the consequences of military aviation noise for both communities and military personnel (Kuehne and Hurley, In Press), studies of how military aviation noise and impacts differ from commercial aviation remain sparse. This research gap becomes particularly critical when the public or agencies seek to evaluate how military operation expansions or alterations may change noise levels, and associated public health impacts, and advocate for mitigation of effects (Kelly, 2019; Tokuda and Barnett, 2017). A fundamental challenge of military aviation has been long-recognized, which is that noise levels are so extreme that “paths of irritation” could extend tens of miles (Bell, 1956; Beranek, 1952); those early predictions have since been confirmed in modeled and empirical studies (Kuehne and Olden, 2020; US Dept of the Navy, 2020). The preponderance of low-frequency noise that propagates readily can result in pervasive human and environmental impacts that manifest across large geographic extents and populations (Jacuzzi et al., 2024; Kuehne et al., 2020; Kuehne and Olden, 2020). Compounding this challenge is that existing assessments of population health consequences are predominantly inferred from studies of commercial aviation, which dramatically differs in aircraft power, speed, elevation, flight frequency, acoustic spectra and loudness (Bell, 1956; Borsky, 1961a; Kuehne et al., 2020; Waitz et al., 2005).

There are substantial logistical challenges associated with the study of public health impacts from military noise. Funding for and incentives to conduct research are limited, in part due to weak policies and regulations for noise assessment and mitigation around military airfields (Appel and Smith, 2011; Kelly, 2019; Kuehne et al., 2020). Access to bases is restricted, operations data are rarely available, and opportunities to coordinate or manipulate flight operations for research are effectively nonexistent. For these reasons, military aviation studies are often opportunistic and occur over limited temporal and spatial scales (Efroymson et al., 2001; Kuehne et al., 2020; Pepper et al., 2003).

Complaint data offer a powerful currency to study the impacts of military noise because they are readily collected by phone, email, and web portals over long time periods and large spatial extents, often with a high level of precision (e.g., automatic timestamps, georeferencing) (Banks and O’Rourke, 2025; Fidell et al., 2012). A further substantial benefit of using complaint data is that, unlike survey-based methods, analysis does not need to be constrained to a limited range of responses that can be assessed within a single study. In noise pollution studies, policy limitations and logistical constraints on study design have resulted in a historical emphasis and nearly exclusive reliance on measurement of annoyance, and to a lesser extent, sleep disturbance, as the primary health outcomes (Clark et al., 2021). Complaint data are inherently unstructured, and thus can reveal the existence and prevalence of a more complete range of impacts (i.e., an individual’s lived experience) to noise and other pollutants on health and quality of life (US Environmental Protection Agency, 1971). While in the past some have considered complaint data as reflecting the opinion of a small number of individuals, substantial research has shown that they have an important and complementary role compared to use of only survey-based methods, which also suffers (albeit less transparently) from respondent bias (Fidell, 2003; Fidell et al., 2012).

Noise complaint data has proven valuable in evaluating impacts and identifying mitigation solutions for aviation noise, dating back to the onset of large-scale commercial aviation and some of the earliest studies of noise from military installations (Bell, 1956; Borsky, 1961a; Mabrey and Carey, 1980). Past approaches have included classifying complaints to describe types of perceived impact (Mabrey and Carey, 1980), using complaint data to identify dominant noise sources in an area (Zambon et al., 2020), examining relationships of complaints to operational and/or acoustic noise characteristics (Hume et al., 2003, 2002), and improving model predictions of noise-induced annoyance or health outcomes (Fidell et al., 2012). More recently, complaint analyses have incorporated automated text-based classifications and emotional content analyses (i.e., sentiment scoring) to reveal patterns and trends in large content environmental datasets, which can then be examined relative to exposure (e.g., air pollution, noise) either spatially or temporally (El Barachi et al., 2021; Fan et al., 2021; Mahl and Guenther, 2023; Wang et al., 2022). All of these methods can be used to create reproducible methods and frameworks for complaint data classification, which can then be applied or modified to analyze comparable datasets (Aiello et al., 2016; Reader et al., 2014; Wang et al., 2022). For example, (Gasco et al., 2020) used crowdsourced social media data to estimate localized noise conditions in London and considerably improve models of predicted health outcomes over conventional noise estimates.

Our study seeks to overcome knowledge gaps with respect to both military aviation and noise assessment generally by analyzing 7 years of compiled complaint data (from two sources) relative to a large military airbase––Naval Air Station Whidbey Island (NASWI) located in Washington, USA. Our study leverages multiple benefits of complaint analysis to assess spatial and temporal trends of community sentiment relative to predicted aircraft noise exposure, and documents the multifaceted ways in which communities are impacted by military aviation noise. Our analyses of the two complaint datasets also allow for assessing patterns over time, including a period before and after a large expansion in training operations. Further, by conducting analyses on two different noise datasets we can assess the reliability of our approaches to reflect the impacts of military aviation on communities, which has historically been a challenging and poorly studied source of environmental noise pollution. The study objectives were threefold: *(1)* characterize the spatial trends in sentiment of complaints, relative to estimated noise exposure; *(2)* evaluate the diversity and prevalence of different types of human health and quality of life impacts for surrounding communities, and; *(3)* identify the operational and environmental factors that drive complaints. These objectives are intended to inform recommendations for operational changes as well as identify areas for future research to assess and mitigate harm to communities exposed to noise from military aviation.

## 2. Methods

The noise complaint data reflect community response to military aviation operations out of Naval Air Station Whidbey Island (NASWI), near the border of Canada and the US in Washington State. Operations are conducted out of two airfields: Ault Field is the primary airfield, with four runways adjoining a naval base; it is located approximately 5 km from the city of Oak Harbor, the largest community in Island County. Outlying Landing Field (OLF) Coupeville is a single runway airfield, located 4 km from the town of Coupeville. The two airfields serve different purposes and have different noise regimes (Jacuzzi et al., 2024). Aircraft are exclusively housed at Ault Field, which serves as the departure point for all training operations as well as interfacility transfers; operations noise associated with Ault Field tends to be consistent but with fewer periods of the intense noise from repetitive training that impacts areas around OLF and its corresponding flight tracks. OLF is used exclusively for “touch-and-go” field carrier landing practice, or FCLPs; these repeated low-altitude (<600 feet) closed loop patterns create intense (>100 decibels or dB) noise during practice sessions. Duration and numbers of practice sessions are highly variable, but can last many hours, including late nights (Jacuzzi et al., 2024; Kuehne et al., 2020).

NASWI was first commissioned and constructed in the early 1940s, with periods of expansion and contraction in operations over time. The most recent and substantial expansion occurred from 2016-2019, with consolidation of the Navy’s fleet of (approximately) 120 electronic warfare aircraft (Boeing EA-18G Growler). The operational expansions resulting from this consolidation were first proposed through two Environmental Impact Statement (EIS) processes under the National Environmental Protection Act (NEPA). The first EIS process - which was limited to the area immediately surrounding Whidbey Island - initiated in 2016 and concluded in 2019 with the Record of Decision, and operations have subsequently increased. This EIS was legally challenged in 2020, resulting in the ultimate finding that the EIS failed to adequately account for several environmental impacts and evaluate alternatives, with an order to revise those components. The EIS was revised, and another public comment and revision period, and the final version filed in December, 2025 (US Dept of the Navy, 2025a). The second EIS [2015 NWTT Final EIS/OEIS], covering a much larger geographic region that includes the Olympic Peninsula, was concluded in 2020.

Since finalization of the EIS’ and resulting Record(s) of Decision, Growler training activities have become widespread in the region, including FCLPs in and around Whidbey Island and electronic warfare and air-to-air combat training in military operations areas on the Olympic Peninsula and in Eastern Washington. Operational flight paths are extensive, and noise from NASWI operations are routinely reported by residents and visitors across at least six counties in Western Washington (Clallam, Jefferson, Island, San Juan, Skagit, and Snohomish), and more intermittently by residents in counties in Eastern Washington. The affected counties are largely ex-urban to rural, with medium to low population density. However, these counties also include areas that are world famous for tourism (e.g., San Juan Islands) and remote wilderness experiences (e.g., Olympic National Park), attracting many millions of visitors each year.

### 2.1. Complaint Data

The two noise complaint distinct datasets were obtained from the NASWI noise complaint line (2017-2020), and Quiet Skies Over San Juan County (2021-2023). Importantly, this period of time encompasses the substantial increase in base operations stemming from the consolidation of Growlers to NASWI. Each dataset is described in detail below.

#### 2.1.1 Naval Air Station Whidbey Island Noise Complaint System

Complaints collected through the NASWI Complaint System were obtained through a Freedom of Information Act Request (FOIA). The purpose of the complaint system is to “assist the installation in maintaining compliance with FAA flight regulations and air station standard operating procedures”(US Dept of the Navy, 2025b). Although personal identifying information was redacted, individuals are given a unique identifier in the complaint database, with repeat complaints assigned to that identifier; this led to identification of 2,324 individuals within the dataset (**Table 1**). Many individuals were classified as “Frequent Callers” (US Dept of the Navy, 2025b); examination of this designation indicated that it was used for callers with approximately two or more complaints within the four-year period, or, in some cases applied to callers with only one complaint. Subsequent communication from the Navy indicated that the threshold for assigning a ‘Frequent Caller’ is the third complaint, regardless of time period (*M. Welding, personal communication*).

**Table 1.** Characteristics of the two noise complaint datasets, which collectively cover a continuous time period of seven years. Location accuracy differed: Quiet Skies complaints are available at high resolution, but redactions during FOIA processing meant that locations of NASWI callers were available to zip code only. Differences in data collection methods mean that not all complaints contain actual comments; subsequent sentiment analyses only utilized complaints that contained comments. The number of unique callers associated with the dataset (across all years) was available by unique identification of the individual within the dataset (NASWI Complaint Line) or was estimated through spatial clustering (Quiet Skies).

| **Datasets** | **NASWI Complaint Line** | **Quiet Skies Website** |
| --- | --- | --- |
| **Years** | 2017-2020 | 2021-2023 |
| **Location accuracy** | Zip Code | Latitude, Longitude (≈ 11 meter spatial resolution based on 4 decimal places) |
| **Number of complaints by year** | 2017: 1,125<br>2018: 871<br>2019: 1,453<br>2020: 3,819 | 2021: 11,070<br>2022: 15,163<br>2023: 14,452 |
| **Number (%) w/ comments** | 6,488 (89%) | 13,784 (34%) |
| **Unique callers<sup>1</sup>** | 2,324 | 50m radius: 8,043 (all complaints)/4,493 (w/comments)<br>100m radius: 5,393 (all complaints)/3,341 (w/comments) |

<sup>1</sup> Unique callers in the Quiet Skies dataset were estimated using the Density-Based Spatial Clustering of Applications with Noise (DBSCAN) algorithm, as described in the Methods.

A known gap in the complaint data was due to unintentional disabling of the email address during the period from August 2020 - February 2021, resulting in loss of any emailed complaints during that time (Stensland, 2021); based on comparison of complaint numbers in the dataset with internal summary reports from FOIA’d documents, the dataset is otherwise assumed to be intact. A sizable percentage of complaints (780 or 11%) contained either brief generic versions of the words “Noise complaint” or were empty (**Table 1**); because complaints are submitted via voicemail message (or email) and transcribed later, it is assumed that these were due to inability to understand or transcribe the comment content.

#### 2.1.2 Quiet Skies Over San Juan County Noise Complaint System

Since 2014, San Juan County and the non-profit organization Quiet Skies Over San Juan County have collected aviation noise complaint data through an ArcSurvey123 reporting system. The website was initiated to document noise issues related to operations from Naval Air Station Whidbey Island, and residents and visitors are “encouraged to use the reporting system whenever they are bothered by jet noise”. Users enter their location, the date and time of the incident, and can leave a 250-character comment; approximately half of complaints that are logged include comments. Prior to 2019, the system was primarily intended to collect noise complaints within San Juan County. In late 2019, Quiet Skies expanded the scope by promoting use of the platform for complaints from any location, and since 2021, out-of-county (i.e., region-wide) complaints have predominated. For this study, we analyzed complaints from 2021 to 2023, focusing on those that included comments.

We used the Density-based Spatial Clustering of Applications with Noise (DBSCAN) algorithm to estimate the number of individuals. This method grouped complaints within a 50-m and 100-m search radius, to approximate the spatial scale of suburban residential lots (Bowen and Geng, 2017); the estimated number of individuals ranged from 3,341 (complaints with comments only, 100-m) to 8,043 (all complaints, 50-m) (**Table 1**). Subsequent analyses, however, were conducted using all complaints with comments, regardless of clustering (i.e., as independent data points). This approach aligns with prior research that analyzed the influence of complaint behavior, and found that high-frequency individuals are legitimate representatives of community sentiment and annoyance (Fidell et al., 2012).

### 2.2 Sentiment and emotion (anger and profanity) analyses

Complaints with free-form comments were analyzed using natural language processing (NLP) and text mining methods. First, the most frequently used words in the comments were computed and illustrated using word clouds, with text size reflecting frequency. Next, exclamation points, often used to express strong emotions, were counted using a custom function to determine their frequency per comment. Additionally, some comments featured text in all capitalized letters, typically representing shouting, yelling, or screaming. A regular expression was applied to identify such words, and their frequency was also visualized using word clouds.

Separate sentiment and emotion analyses were conducted to evaluate overall sentiment, expressions of anger, and profanity. The comments were first tokenized into sentences, and words were matched to a predefined lexicon of polarized terms using the *sentimentr* package in R (Rinker, 2021a). Negative polarity words such as “loud,” “noise,” and “disrupt,” as well as positive polarity words like “peace,” “quiet,” and “good,” were identified. Valence terms that modify sentiment (e.g., “never,” “however,” “although”), were also taken into account. The sentiment score for each comment was derived by summing the weighted values of positive and negative words, adjusted by valence shifters, and averaging across the sentence level, resulting in an overall sentiment score that categorizes comments as negative, neutral, or positive.

As complaints are generally expected to convey negative sentiment, further analysis was conducted to identify expressions of anger using a lexicon of anger-related words (Rinker, 2021b). Common anger words such as “scream,” “noisy,” and “horrible” were identified, and an anger score was calculated as the rate of anger words per comment. Profanity was analyzed using the same lexicon, and a score was calculated as the rate of profanity-related words per comment.

Since the lexicon used does not include grawlix (i.e., strings of typographical symbols such as “#@*%” used in place of profanity), a regular expression was used to identify comments containing at least four consecutive typographical symbols as grawlix. While grawlix occurrences were not included in the profanity score calculation, the words surrounding grawlix in the comments were visualized with a word cloud.

Kernel density maps were generated to illustrate the spatial distribution of complaints with the most negative sentiment, highest anger scores, and highest profanity levels. For the sentiment analysis, only complaints with negative scores were included. Lexicon-based sentiment methods cannot reliably distinguish sarcasm or rhetorical expressions, which may occasionally produce positive sentiment despite clearly negative intent. To avoid misclassification and to conservatively emphasize areas of concern, positive sentiment scores were set to zero rather than treated as evidence of positive sentiment. Each point was weighted by the product of the number of complaints and the average score (sentiment, anger, or profanity) associated with the location:

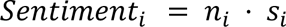

where *n_i_* is the number of complaints and *S_i_* is the average score at location *i*. The resulting kernel density surfaces reflect the spatial intensity of community response based on the magnitude and the concentration of complaints. To better understand the geographic distribution of complaints and associated emotional responses, we aggregated complaint data spatially and overlaid these results with modeled noise exposure contours (L_dn_, day-night average sound level) from military flight operations generated by a previous study. Briefly, the noise exposure contours are generated from a standard U.S. Department of Defense simulation model used by the Navy to predict noise exposure from flight activity (Jacuzzi et al., 2024).

All text analysis was performed in R v4.2.3 (R Core Team, 2025), using the tidytext v0.3.4 (Silge and Robinson, 2016), sentimentr v2.9.0 (Rinker, 2021a), SnowballC v0.7.0 (Bouchet-Valat et al., 2023), stringr v1.5.0 (Hadley, 2023), and ggplot2 v3.4.1 packages (Wickham, 2016). Cluster analysis was conducted using the dbscan package v1.1.12 (Hahsler et al., 2023).

### 2.3 Taxonomy of noise impacts and key term identification

To evaluate and compare the prevalence of different types of impacts from jet noise, it was first necessary to develop a taxonomy of categories and subcategories of impact. We initiated this taxonomy by reviewing the diverse impacts of noise that are reported in the literature for health outcomes and quality of life, including in wilderness or natural areas (Basner et al., 2014; Borsky, 1961a, 1961b; Buxton et al., 2019; Mace et al., 1999; US Environmental Protection Agency, 1971; Wemheuer, 1979; World Health Organization, 2011). Based on our review of both the literature and the complaint data, we identified 22 distinct types of potential impacts, which we organized into seven broad categories: Work, Recreation & Social, Health, Home Experience, Children, Life Quality, and Environmental Impact. Each category is composed of 2-4 subcategories that specify distinct aspects of the broad category (**Table S1**). Following (Montini et al., 2008), we also included a broader contextual category of Operations (with two subcategories) to capture when complaints related to the military base, flight patterns, or specific flight events.

Using this taxonomy and knowledge from the complaint content, we identified keywords uniquely associated with each subcategory and were not highly contextual (i.e., the word would be unlikely to indicate a different category depending on context). This initial keyword set was used to classify complaints, from which sets of 100 randomly selected classifications were evaluated, and the keyword sets updated iteratively to reduce errors of omission and commission. Omission rate was reduced by addition of unique keywords, and commission by the removal of some words that proved to be contextual (e.g., ‘tired’ could indicate lack of sleep but also annoyance, as in “tired of noise”) and/or use of word pairs for greater specificity. Error rates were reduced to 9% and 8% for omissions and commissions, respectively, which was determined based on a final validation of 500 randomly selected complaints. The validation process also confirmed that complaints that did not classify were more general in nature or described an impact that was not part of the taxonomy. Lastly, the validation process resulted in the combining of one initial subcategory with very few keywords (i.e., “Disruption of learning at home”) into a similar subcategory (“Disruption of Learning”), for the final 21-item taxonomy; the final taxonomy and keyword sets are found in **Table S1**.

### 2.4 Modeling factors associated with daily complaints

We used generalized linear modeling (GLMs) to identify the effect of operational and environmental factors on the number of complaints each day. GLMs are an extension of simple linear models that accommodate response variables with different distributions (e.g., count data). Specifically, our analysis leveraged hurdle models, two-part models to account for zero-inflation, and where zeros are likely to stem from different mechanisms than positive counts (i.e., structural zeros) (Hu et al., 2011). In our datasets, zero complaints were more likely on days with no scheduled FCLPs (e.g., weekends, other periods of no operations). The count model for complaints was fit using a negative binomial distribution and log link. The zero-inflated component (i.e., where days with non-zero complaints were predicted by a binary variable of scheduled FCLP operations) was fit using a binomial model and a logit link. For the negative binomial count model, the dispersion parameters were tested for difference from zero with t-statistics.

The operational variables included in models were derived from a dataset of Field-Carrier-Landing-Practice (FCLP) daily schedule notifications, which are distributed on a weekly basis by the Navy. The schedule is distributed as a courtesy and subject to change, but is the only available indicator of operations, and has shown to correspond with measured noise from operations in other analyses (Kuehne et al., 2020; Kuehne and Olden, 2020). We filtered the FCLP data to only include operations scheduled for the Outlying Field Coupeville (OLF) airfield, and correspondingly filtered the noise complaints in both datasets to zip codes that were most likely to be impacted by OLF flight paths. This was due to the fact FCLPs constitute the primary/sole operations at OLF, and complaints originating around OLF are therefore more likely to be in response to those operations. In contrast, complaints originating in proximity to Ault Field (e.g., Lopez Island, LaConner) could be in reference to FCLPs but are more likely to be in response to other operations (e.g., runups, carrier transits, other aviation exercises) with unknown schedules (Jacuzzi et al., 2024). The zip codes nearest to and surrounding OLF included the cities of Coupeville, Oak Harbor, Camano Island, Langley, and Freeland in Island County, and Port Townsend in Jefferson County.

The FCLP schedule is published using eight timeframes, which we collapsed into a daily count of FCLP sessions for three periods: Day (timeframes = Early/Late Morning, Early/Late Afternoon), Night (timeframes = Early/Late Evening, Night), and Late Night (timeframes = Late Night, Past Midnight). To test for a cumulative effect of operations on the number of complaints, we also calculated lagged operational variables, corresponding to the summed FCLP sessions for 1-6 days, 14 days, and 30 days prior. Daily temperature and precipitation (measured at nearby Seattle-Tacoma International Airport) were included as variables in candidate models, as indicators for the opportunity to spend time outdoors, as well as the potential need to keep windows and doors open in summer for cooling. Lastly, season (winter, spring, summer, autumn) and year were included in candidate models to identify differences in complaints that could be associated with seasonality or in specific years. Variables were tested for multicollinearity; the 1-6 day cumulative session variables were strongly correlated, and were therefore included only in separate candidate models.

AIC model selection was used to distinguish among candidate models (Pinheiro and Bates, 2000), and indicated that the model with all parameters was the best-fitting for both the Navy Line and Quiet Skies datasets. All data summaries and statistical analyses were conducted in the R Statistical Programming Environment (R Core Team, 2025).

## 3. Results

### 3.1 Spatial and Temporal Patterns of Complaints (Quiet Skies)

A total of 11,070, 15,163, and 14,452 complaints were submitted to the complaint site in 2021, 2022, and 2023, respectively. Of these, 4,937, 5,028, and 3,819 included comments for each year (**Table 1**). General temporal patterns of complaints across the years were aligned with military aviation operations from NASWI: complaints occur year-round, most prominently during daytime to evening hours when the largest number of FCLP and other operations are scheduled (**Figures 1 and 2**, **Table S2**). A large majority of complaints are made on weekdays, which is consistent with a preponderance of flight operations on weekdays, both around NASWI (Jacuzzi et al., 2024) and the Olympic Peninsula (Kuehne and Olden, 2020). A word cloud of the highest-frequency words is shown in **Figure S1**, with prominent terms including “growler”, “loud”, “noise”, “jet”, and “fly”.

**Figure 1.**
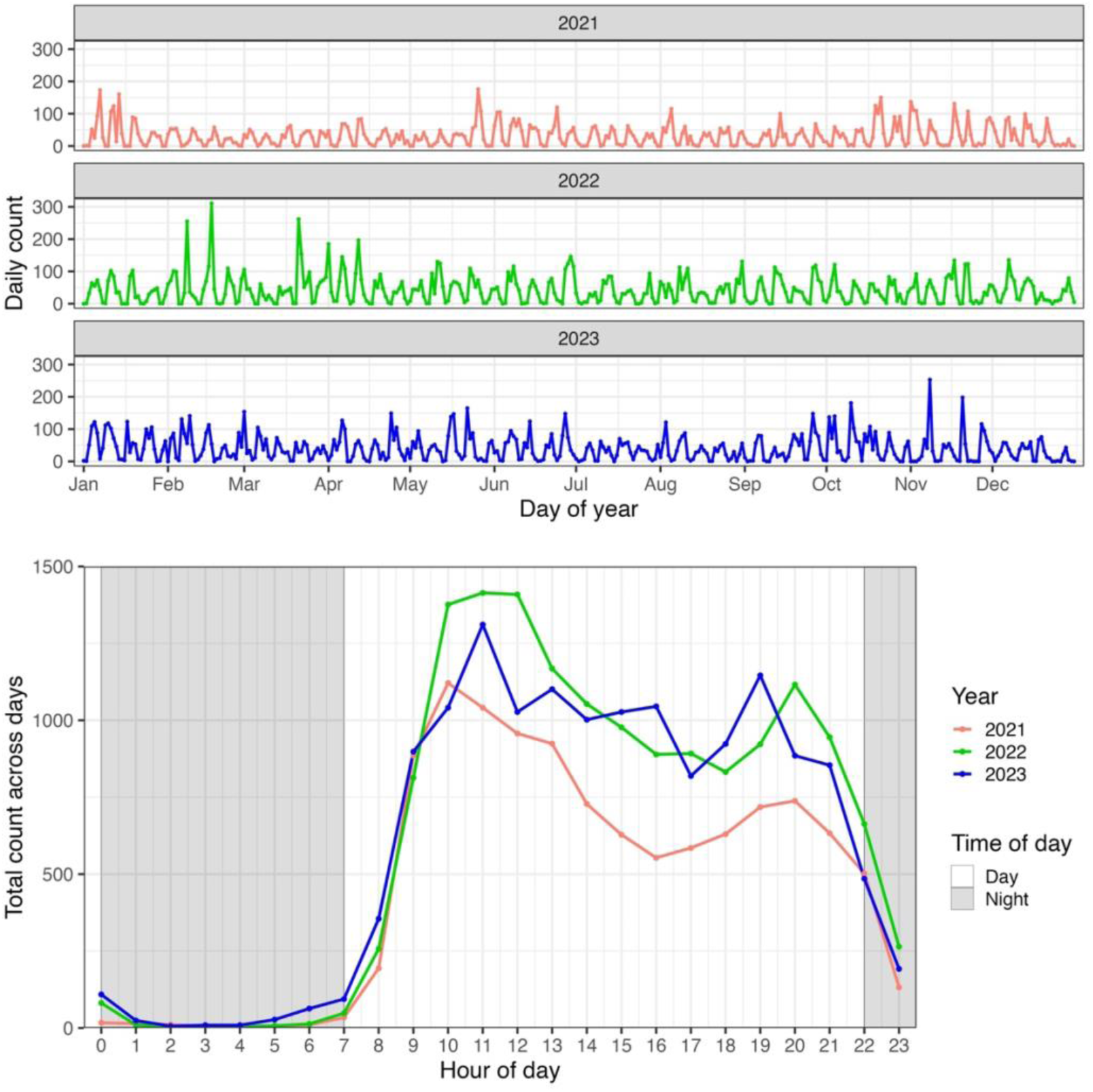
Temporal distribution of Quiet Skies noise complaint (2017 - 2020). Top: Daily complaint counts by year. Bottom: Hourly distribution of complaints, aggregated across all days.

**Figure 2.**
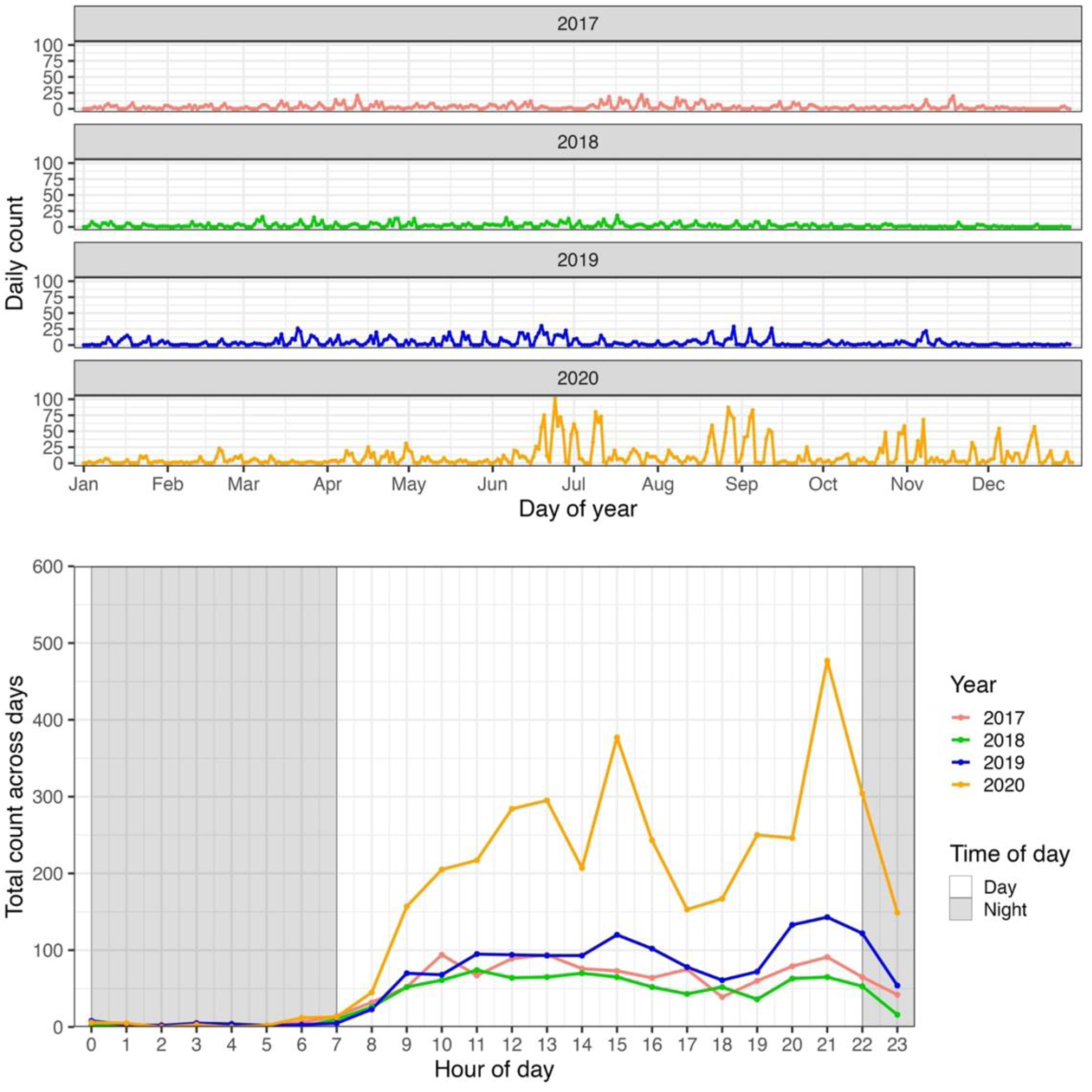
Temporal distribution of Navy line noise complaint (2017 - 2020). Top: Daily complaint counts by year. Bottom: Hourly distribution of complaints, aggregated across all days.

### 3.2 Sentiment, anger, and profanity in complaints

As expected for complaints, the average comment sentiment was negative and remained consistent for all years. The mean sentiment score (SD) from comments were -0.17 (0.26), -0.16 (0.26), and -0.17 (0.25) for 2021, 2022, and 2023, respectively. As an example, in 2022, the words most frequently associated with the comments that scored in the top 25 percentile for negative sentiment were “noise” (n = 559), “Growler” (n = 273), “loud” (n = 215), “jet” (n = 151), “day” (n = 84) and “vibrate” (n = 84). Of these 5,032 comments, 1,462 scored positively for anger emotion.

The mean anger score (SD) in year 2021, 2022, and 2023 were 0.028 (0.08), 0.027 (0.07), and 0.024 (0.06), respectively. Comments consisting of a single word that was also identified as an anger word received the highest anger score (1.0), e.g., *“Bad!!!”, “Brutal”, “HORRIBLE!!!!!!”, “Menacing!!!”.* For comments with more than one word, high-scoring examples included: *“Damn noisy growlers making life hell below”, “Blast after blast of rolling sound”,* and *“Extremely oppressive roaring.”*

The proportion of comments containing profane words has increased over the years. In 2021, 2022, and 2023, profane words appeared in 2%, 3%, and 10% of all comments, respectively, with corresponding mean profanity scores (SD) of 0.005 (0.04), 0.004 (0.04), and 0.011 (0.04). Use of profanity in comments reflect the profound impacts of noise on individuals and families in the region, the repeated and frequent occurrence of noise, and the blame placed on the Navy, and a perspective that the Navy is permitted to create noise that others are not allowed. For example, “*Again! This is so f***ing ridiculous. Today was the first day of school and my kids are still fucking awake because of these jets. How is this allowed? Why is this not scheduled better? Really? At 11pm? Are you f***ing kidding me?”* (profane words are censored).

Across all years, some comments contained grawlix, indicating the presence of profanity. These comments—whether using symbols or explicit profane language—often expressed similar concerns, particularly regarding the intensity and frequency of the noise and its connection to naval activities. For example: “*Memorial Day. South Lopez. Sounds like we will get the fall out jet noise from Navy’s “flyover” of Oak Harbor. SCREAMING JETS OVER US NOW. GO AWAY. &*!!&^%$$#%^&%$ Pilots just having fun while we are abused. Waste of money.*” The most common words used in combination with profanity/grawlix were “jet” (N=145), “noise” (N=134), “loud” (N=130), “fly” (N=124), and “lopez” (N=103). These words were also among the most used words in comments in general. Other terms indicating noise source, e.g., “Navy” (N=60) were included amongst these comments. Terms indicating health impacts were also mentioned, e.g., “sleep” (N=56) and “disrupt” (N=27).

Complaints included the use of capitalized words as a way of yelling or shouting to express strong feelings through a typographical tone of voice. Some of the capitalized words represent acronyms (e.g., OLF, NAS, US, and PM). However, the most frequently yelled words also include noise descriptors such as “loud (N=562)”, “roaring (N=188)”, “screaming (N=74)”, “another (N=178)”, “overhead (N=71)”, and “growler (N=71)”. Similarly, exclamation marks were used as a typographic indicator of intensity, with 2,034 complaints containing at least one, and 214 containing three or more. The most extreme example included 153 exclamation marks.

The spatial variation in negative sentiment scores shows pronounced clusters of more negative sentiment in areas with higher complaint volumes and close proximity to high levels of noise exposure (**Figure 3**). As illustrated in **Figure S2**, the density of complaint reports, with the highest concentrations observed in areas located under or adjacent to documented flight paths, particularly over Whidbey Island, Camano Island, and extending into parts of Jefferson, Island, and San Juan Counties. **Figures S3 and S4** similarly show the spatial distribution of anger and profanity intensity scores, respectively. While many of the highest intensity values align with areas of modeled elevated noise exposure, we also observed elevated emotional responses and substantial complaint activity in areas outside the modeled contours, including parts of the San Juan Islands and more distant locations.

**Figure 3.**
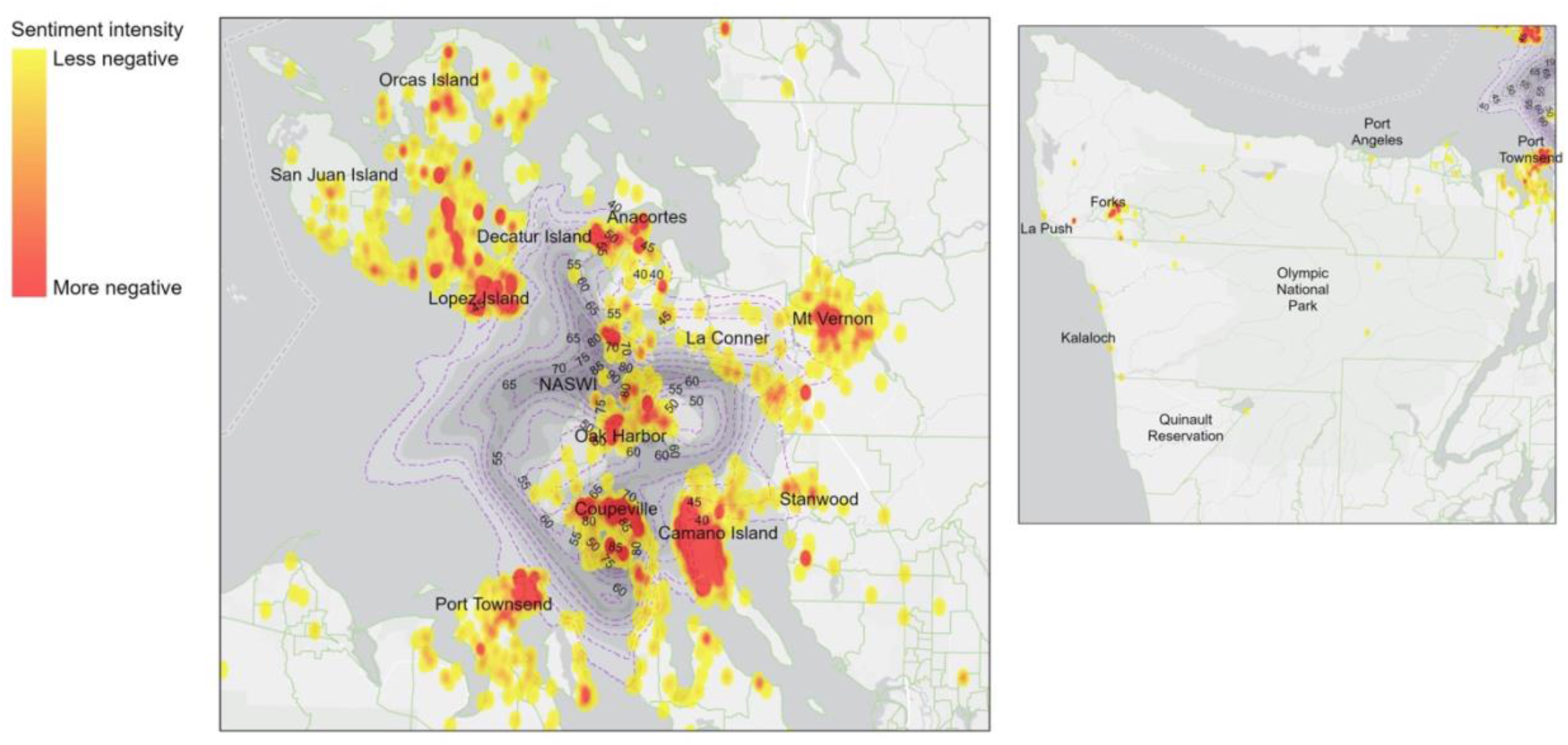
Kernel density of negative sentiment intensity in complaints, overlaid with modeled day-night noise level (L*_dn_* dBA). Left: NASWI Whidbey Island vicinity. Right: Olympic Peninsula.

### 3.3 Noise impacts on health and well-being

Using the final taxonomy and lexicon, 55% of Navy line complaints and 48% of Quiet Skies complaints were classified into one or more categories of impact (**Figure 4a-b**). Of the complaints that were classified, more than half of those (56%) were to a single category, with declining percentages up to a maximum of 9 categories of impact referenced in an individual complaint. Both datasets showed similar patterns in the representation of different categories (**Figure 4a-b**): Home Enjoyment & Use was consistently a subject of complaint content, with frequent mentions of Home Use, Home Damage (e.g., vibration and shaking), and effects on Pets & Domestic Animals. Health Concerns was the next most common category of complaints, within which Sleep Disruption was the most prevalent, followed by Hearing Concern.

**Figure 4.**
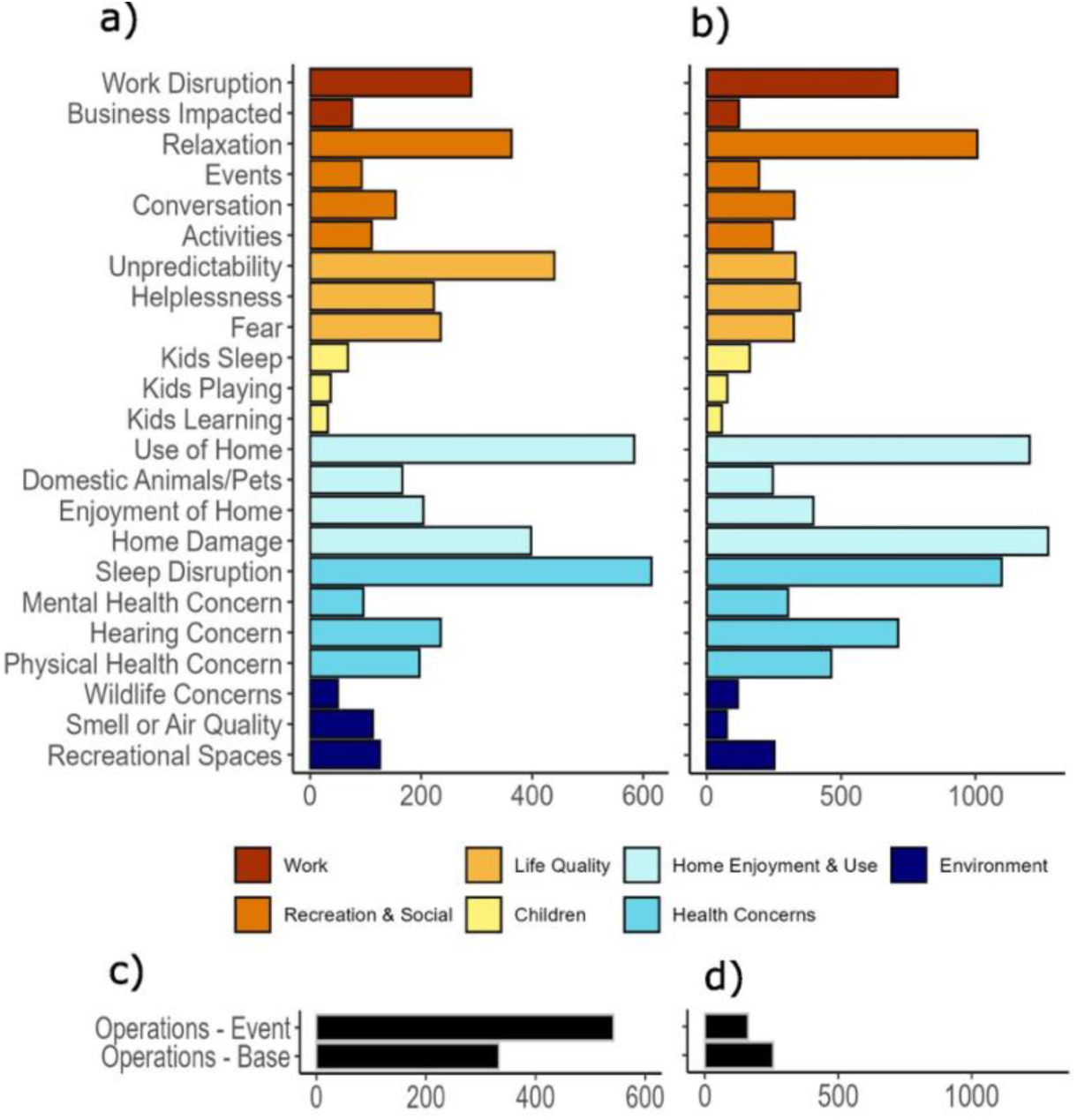
Classification of a) Navy line (*n*=2,022) and b) Quiet Skies (*n*=4,073) complaints into 23 subcategories within seven broad categories (colored bars) describing impacts on health and quality of life and c-d) complaints related to specific flight events and/or general base operations.

Complaints related to Recreation & Social Activities were common; in particular, the Relaxation subcategory was prominent - which would include activities like reading, watching TV, or listening to music. Complaints also commonly referenced impacts on Work. Importantly, all three subcategories within Life Quality (Unpredictability, Helplessness, and Fear) were consistently featured, indicating a high degree of frustration and distress in the community. The primary distinction between the two datasets lies in the category of Unpredictability (**Figure 4a-b**) and in the Operations category (**Figure 4c-d**), where complaints to the Navy line were more likely to reference concerns about schedule of operations, flight path and patterns, a particular flight event or the military base.

The representation of different complaint categories varied in different locales (**Figures S5 and S6**), consistent with differing proximity to the airfields and the types and numbers of flight operations that residents experience, based on their location. Complaints from residents of Island County (i.e., cities of Coupeville and Oak Harbor) reflect high levels of impact across all complaint categories, while Jefferson County (i.e., Port Townsend) and San Juan Island communities - located farther from the airfields - reflect fewer categories and level of impacts overall. Notably, complaints from residents in Clallam County (i.e., Olympic Peninsula, Forks) reflect a large and similar range of impacts, despite differences in the nature of the operations (i.e., electronic warfare and air-to-air combat training). Despite differences in number of complaints across locations, the categories of Home Enjoyment & Use and Sleep Disturbance were consistently featured, suggesting that these categories of impact are among the most problematic for residents.

### 3.4 Factors associated with daily complaints

Our modeling of operational and environmental factors driving daily complaints resulted in good estimates of model fit (*dispersion parameter* = 1.01 for Navy Line, 0.97 for Quiet Skies) and high *R^2^*(**Tables 2-3**). Within the zero-inflated (i.e., hurdle) portion of the models, the baseline odds of having positive (i.e., non-zero) complaints on any day differed somewhat between the two datasets, with a reduced odds (*OR* = 0.79, 95% CI [0.70 – 0.89]) for the Navy Line dataset, and a higher baseline odds (*OR* = 1.21, 95% CI [1.05 – 1.40]) of non-zero complaints within Quiet Skies data (**Tables 2-3**). These odds increased dramatically and significantly on days with scheduled FCLP sessions: scheduled operations increased the odds of complaints by 13.8 times (CI [9.40 – 20.37]) in the Navy Line, and by 23.5 times (CI [12.99 – 42.57]) in the Quiet Skies data.

**Table 2.** Generalized linear model results for complaints from the Navy line dataset, filtered for the zip codes surrounding the Coupeville Outlying Field, for the years 2017-2020. Incidence rate ratios show the effect size of operational and environmental variables on the number of complaints each day. Operations variables are based on the number of scheduled Field Carrier Landing Practices (FCLPs) for different timeframes (Day, Night, and Late Night) as well as the 1-day and 30-days prior.

| Predictors | Effects of Variables on Complaints |  |  |
| --- | --- | --- | --- |
|  | Incidence Rate Ratios | CI | p |
| **Count Model** |  |  |  |
| (Intercept) | 0.59 | 0.33 – 1.06 | 0.077 |
| Season [Spring] | 0.98 | 0.75 – 1.27 | 0.869 |
| Season [Summer] | 1.03 | 0.73 – 1.46 | 0.866 |
| Season [Autumn] | 0.90 | 0.68 – 1.19 | 0.443 |
| Year [2018] | 0.89 | 0.69 – 1.13 | 0.333 |
| Year [2019] | 1.11 | 0.86 – 1.43 | 0.435 |
| Year [2020] | 2.61 | 2.06 – 3.30 | **<0.001** |
| Day FCLPs | 1.54 | 1.39 – 1.70 | **<0.001** |
| Night FCLPs | 1.35 | 1.20 – 1.50 | **<0.001** |
| Late Night FCLPs | 2.41 | 1.95 – 2.96 | **<0.001** |
| Prior FCLPs – 1 Day | 1.12 | 1.05 – 1.18 | **<0.001** |
| Prior FCLPs – 30 Day | 0.99 | 0.99 – 1.00 | **0.003** |
| Temperature | 1.02 | 1.01 – 1.03 | **0.002** |
| Precipitation | 0.57 | 0.38 – 0.87 | **0.009** |
| **Zero-Inflated Model** |  |  |  |
| (Intercept) | 0.79 | 0.70 – 0.89 | **<0.001** |
| Scheduled FCLPs [Y] | 13.83 | 9.40 – 20.37 | **<0.001** |
| **Observations** | 1461 |  |  |
| **R<sup>2</sup> / R<sup>2</sup> adjusted** | 0.980 / 0.979 |  |  |

**Table 3.**
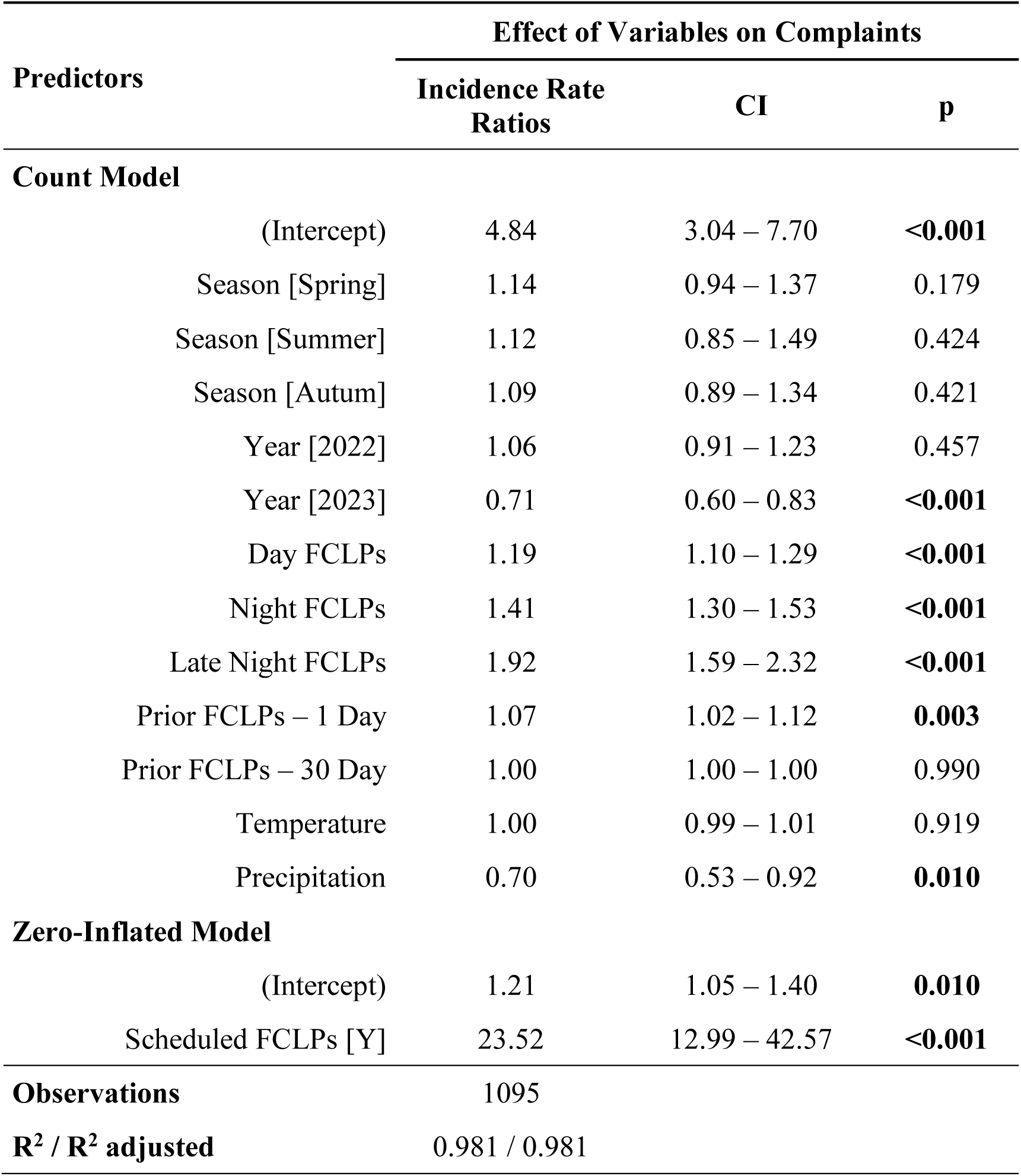
Generalized linear model results for complaints from the Quiet Skies dataset, filtered for the zip codes surrounding the Coupeville Outlying Field, for the years 2021-2023. Incidence rate ratios show the effect size of operational and environmental variables on the number of complaints each day. Operations variables are based on the number of scheduled Field Carrier Landing Practices (FCLPs) for different timeframes (Day, Night, and Late Night) as well as the 1-day and 30-days prior.

Within the count model (i.e., non-zero complaint days), the number of FCLP sessions scheduled in the three time periods - Day, Night, and Late Night - significantly increased the incidence rate, for both datasets (**Tables 2-3**). In the Navy Line model, each unit increase (i.e., added FCLP session) during the Day increased complaints by 1.54 times, at Night by 1.35 times, and during Late Night by 2.41 times. Each additional FCLP session 1-day prior increased the incidence rate of complaints by 1.12 times, but the cumulative sessions in the 30-days prior were associated with a very slight but significant decline (*IRR*=0.99). Higher temperatures were associated with a slight but significant increase in complaints (*IRR* = 1.02), where higher precipitation was associated with a substantial and significant reduction in numbers of complaints (*IRR* = 0.57).

These patterns were largely mirrored in the Quiet Skies dataset. FCLP sessions during the Day increased complaints by 1.19 times, at Night by 1.41 times, and during Late Night by 1.92 times. Each additional FCLP session the 1-day prior increased complaints by 1.07 times, but there was no effect of sessions in the 30-days prior. In the Quiet Skies model there was no significant effect of temperature (*IRR* = 1.0), but higher precipitation was still significantly and strongly related with reduced complaints (*IRR*=0.70).

Season had a relatively neutral effect in both models (**Tables 2-3**), which is consistent with the year-round nature of operations at WINAS. The effect of Year was stronger, particularly in the Navy model, when complaints increased by 2.6 times in 2020, compared to 2017 (the intercept) (**Table 2**). The year 2020 was important in being both the first year of increased operations following the 2019 Record of Decision, and the initiation of COVID-19 pandemic and lockdowns. In the Quiet Skies model, the effect of year was seen in a significant decline in complaints in 2023 (*IRR* = 0.71), compared to 2021 (the intercept) (**Table 3**).

## 4. Discussion

In this study, we sought to develop and apply new methods to the analysis of complaint data to quantify the impacts of noise exposure in communities, particularly those that are impacted by poorly studied military aviation noise. We used sentiment analysis to map and test correspondence of negative sentiment with modeled noise exposure from Naval Air Station Whidbey Island (Washington, USA), developed a taxonomy and lexicon to identify the diversity and prevalence of impacts on health and quality of life, and modeled the dominant operational and environmental factors driving complaints. In accomplishing these objectives, our study demonstrated an abundance of information available in noise complaints, which are commonly available data and readily collected in noise-impacted communities. Given the dearth of research and challenges in studying noise impacts from modern military aviation and airbases, our study also represents a substantial advancement to develop informed monitoring, assessment, and mitigation recommendations.

Our analysis of sentiment, anger, and profanity in complaints provides a comprehensive framework for assessing the volume and motivation of complaints as well as the emotional intensity of community responses to military aviation noise. This assessment highlights not only the frequency and nature of disturbances but also captures their psychological and emotional toll. The distinction between short-term reactions (complaints) and longer-term stable responses (annoyance), as noted by Fidell and Howe (1998), underscores the importance of integrating qualitative emotional metrics to complement traditional measures of noise impact. By analyzing emotional responses, this study captures a richer understanding of the lived experiences of affected residents.

Contrary to the assumption that complaints are driven by a small subset of frequent complainers, our results demonstrate widespread impacts, with intense negative sentiment and anger distributed across a large geographic area surrounding NAWSI. Areas closest to airfields and flight paths, such as Coupeville and Forks, consistently exhibited the most intense negative sentiment and anger. However, areas outside estimated noise contours, such as parts of the San Juan Islands, also exhibited high levels of negative sentiment and anger. These results are consistent with other studies that have shown a high degree of annoyance and complaints can occur at lower levels of annual noise exposure (Fidell and Howe, 1998), particularly in response to military aviation noise regimes of high intensity (Banks and O’Rourke, 2025; Gelderblom et al., 2014; Yokoshima et al., 2021). Our results support a growing body of evidence highlighting the importance of renewed approaches to account for event-specific impacts in addition to cumulative impacts, which remain absent from noise assessment guidelines currently employed worldwide. Relatedly, factors such as predictability and perceived control, which Guski et al. (1999) identified as significant in shaping annoyance responses, likely contribute to the spatial variability observed in our results. These findings point to potential shortcomings of traditional noise indicator metrics (i.e., A-weighted decibels) and modeling procedures (i.e., annualized noise exposures) to accurately predict the response experienced by exposed communities. Incorporating spatially explicit analyses of emotional responses offers an opportunity to refine models and improve our understanding of how noise affects communities.

The strong association between military aviation noise and overall negative sentiment, anger, and profanity reflects a cumulative emotional and psychological toll of noise exposure from training operations. Persistent and intense exposures drive heightened expressions of frustration and distress, as evidenced by the prevalence of emotionally charged language, typographical emphasis, and the increasing use of profanity over time. These findings are consistent with (Stansfeld and Matheson, 2003), who emphasized the role of chronic noise exposure in inducing stress and negative emotional states, particularly in contexts of prolonged or unpredictable noise events.

The ability to map sentiment, anger, and profanity scores provides actionable insights into the geographic distribution of community impacts. These maps highlight hotspots of distress near airfields and flight paths, while also revealing unexpected patterns in areas outside estimated noise contours. These unexpected patterns are primarily high density and intensity of complaints at the periphery or even entirely outside of the modeled contour lines, including areas of Lopez Island, Camano Island, and the towns of Mount Vernon, and Anacortes. Fidell and Howe (1998) emphasized the value of spatial analysis in understanding annoyance, noting that noise complaints provide unique opportunities for uncovering temporal and spatial patterns of community response. By incorporating spatially explicit emotional metrics, this approach provides a valuable tool for identifying priority areas for intervention and guiding policy decisions to reduce community burdens.

A further strength of utilizing complaint data is the ability to evaluate a broad range of ways in which noise can impact people and communities. These complaint data were collected over long time periods, allowing us to identify consistent patterns across two different datasets. Currently, at a global scale and across all forms of transportation noise, only a handful of noise impacts ––typically annoyance, followed by sleep disturbance –– have been studied robustly enough to allow quantitative prediction of population health impacts (World Health Organization, 2022). However, policy implementation of measures to reduce even these impacts can vary greatly across jurisdictions and geography (Jacuzzi et al., 2024). Studies of military aviation noise have documented population health impacts ranging from annoyance to sleep disturbance and a suite of physiological health outcomes including increased anxiety and depression, prevalence of hypertension or cardiovascular impacts, and hearing concern or loss (Ising et al., 1990; Kim et al., 2014; Okinawa Prefectural Government, 1999; Olden et al., 2026; Yokoshima et al., 2021). Our analysis indicated that a majority of these impacts are being experienced in communities, and are reflected to some extent in complaint data. Importantly, our analysis indicates the prevalence of different impacts on communities. These include inability to plan work or social events, reduced value or enjoyment of home or neighborhood, reduced value of natural and recreation areas, and impacts on family life and pets. Our analysis also reveals a range of more extreme impacts, where complaints reference concerns about hearing damage or loss, vibration and/or damage to homes, fear, helplessness, and affects on childhood learning and well-being.

Our analysis can also be used to guide and prioritize mitigation opportunities, for NASWI and for other airfields. Many military airbases maintain complaint data systems (Mabry and Carey, 1980). By developing a comprehensive taxonomy of impacts and a lexicon for mapping complaint content to impacts, this approach can be used to evaluate how a community is responding to operations and indicate what approaches may best offer relief. For example, we found that a large number of complaints related to unpredictability with respect to the published schedule, and corresponding inability to plan work, time outdoors, or social events, while others reference unpredictability in spatial patterns or flight paths being flown. These findings suggest that a commitment by NASWI to scheduling, communicating, and adhering to regular training schedules and flight paths would be an important step in helping relieve the public health burden on communities. Other prevalent categories related to compromised use or enjoyment of homes and neighborhoods, including frequent references to vibration and shaking of windows, walls, and home contents and/or using hearing protection inside homes. Given the extreme degree of noise exposure experienced by some residents on a routine basis, mitigation strategies including soundproofing and property purchases should be fully considered and explored. While these policies are implemented by military airfields in other countries (Tsukioka et al., 2011; Weinandy et al., 2014), current US domestic policy to mitigate noise around airfields is limited to advising localities on land use (Appel and Smith, 2011).

Modeling of the factors driving the daily number of complaints were extraordinarily consistent, even given differences in the purpose and time period of the two datasets. We found that the number of complaints was strongly related to the number of (published) operations that day (Hume et al., 2003). This result is important for two reasons. First, the schedule of FCLP operations is not an official schedule and is published as a courtesy, being subject to change without notice and based on general daily timeframes. That we detected a strong relationship even with this imperfect predictor suggests the relationship would be even stronger with a more accurate indicator of operations. Second, a common interpretation of noise complaint systems and reporting is to ascribe complaints to a handful of “frequent callers” (Fidell et al., 2012; Fidell and Howe, 1998), a designation that is assigned to a caller on their third complaint, regardless of the period of time (i.e., many years) when the complaints were made. We found in fact that 90% of callers to the Navy Line submitted less than 5 complaints over a 4-year period, and our results demonstrate that these complaints are in fact responsive to the number and timing of operations and/or specific flight events (van Wiechen et al., 2002). We also found a cumulative effect, where there was a consistent effect of operations on the previous day. This trend reflects a fairly common sentiment in the data, that many people wait for some time or reach some threshold of frustration, before making a complaint. Lastly, our analysis demonstrates the highly disproportionate influence of late night operations on the number of complaints (Hume et al., 2003). These results suggest that mitigation and relief to communities should prioritize reduction or cancellation of late night operations. More spacing between training sessions and operations could also reduce the cumulative effect that we observed, especially in combination with communication of accurate schedules to reduce unpredictability.

As has been found in other studies, we did find evidence for relationships of complaints with environmental conditions (Dreier and Vorländer, 2021; Fidell and Howe, 1998). We found a small but consistent positive relationship with temperature, and a negative effect of precipitation. The strongest relationship with temporal factors, however, was the large increase in the summer and autumn of 2020. This period corresponded with both an increase in operations following the 2019 Record of Decision and with the onset of the COVID-19 pandemic and lockdowns.

An important result of the spatial mapping of complaints - both the numbers and the intensity of negative sentiment - is demonstrating patterns of community frustration relative to the modeled exposure levels. Our results revealed not only high negative sentiment in close proximity to the airfields and flight paths, but also in areas that are estimated as being minimally or even unimpacted based on annual modeled noise. These noise contours represent day-night average sound levels (L_dn_) at an annual scale, generated from a standard United States Department of Defense simulation model used by the Navy to predict noise exposure from flight activity (Jacuzzi et al., 2024). The conventional practice of predicting exposure from annual cumulative metrics has been widely criticized as being inadequate and inappropriate to assess the impact of periodic, intermittent, high-intensity noise from military aviation (Gelderblom et al., 2014; Jacuzzi et al., 2024; Kerry et al., 1998; Yokoshima et al., 2021).

Periodicity or intermittency of flights is challenging to study, and to date, the only research on how this relates to annoyance to military aviation was a single study in the 1980s that tested a small number of flight events per day (min-max: 6-20, mean: 12), only half of which were high noise exposure (maximum outdoor sound exposure level of 110-115 dB) (Stusnick et al., 1993). Operations at NASWI can be well over 10-fold this number in a single day, extending over many hours, and at higher received noise levels (Jacuzzi et al., 2024; Kuehne and Olden, 2020).

Operations on the Olympic Peninsula can be 6-fold this number of highly variable overflight events ranging from a few seconds to more than twenty minutes at a time, and resulting in hours of cumulative exposure (Jacuzzi et al., 2024; Kuehne and Olden, 2020). We suggest that research is needed to establish accurate understanding of how current levels and numbers of high-intensity, intermittent flight events are impacting communities, to inform development of noise modeling that reflects realized impacts and lived experience (Banks and O’Rourke, 2025).

Our analyses indicate short-term priorities to address challenges of military aviation noise. Among the most impactful findings is the clear and disproportionate influence of late-night operations on complaint volume and emotional intensity, suggesting a clear priority for operational mitigation. Our results also demonstrate intense, consistent frustration with the unpredictability of flight schedules and locations, which could potentially be mitigated by more transparent and consistent communication between military authorities and affected communities. The sentiment analyses revealed an apparent strong mismatch between noise exposure estimated by noise modeling, and the noise exposure and annoyance experienced in the community. Comprehensive research to better understand the reasons for this misalignment (e.g., infrequent but high-intensity operations, reliance on annual operations data in models) could indicate how noise modeling may need to be adjusted to capture community impacts. The geographic placement of NASWI, which is adjacent to communities with high proportions of recreational users and seasonal residents, may also be exacerbating negative sentiment, as these groups have expectations of a quiet environment. A better understanding of how military aviation is affecting not only residents but recreational area users could help explain widespread negative sentiments.

Because many areas that experience noise from NASWI are not within the modeled noise contours, it is difficult to estimate with precision the percentage of the affected population that actually submitted complaints. However, using the estimate of 74,000 people in the region exposed to noise levels associated with “adverse effects” (Jacuzzi et al., 2024), our results indicate that approximately 3-10% of the population are motivated to submit at least one complaint within a 3-4 year period (alternatively, 2-6% of the combined population of the three most impacted counties). Analysis of complaints for civil aviation has reported that no more than 6% of a population, even if “greatly bothered”, will express disagreement or complaint (Hart et al., 1954). The relatively high level of complaint behavior around NASWI indicates widespread frustration and the likelihood of ongoing community resistance and legal actions (Wemheuer, 1979).

Our results also offer insights toward needed development of research and policies to mitigate military aviation noise at the federal level, to help address the longstanding challenges at NASWI and other air bases and airports, including deployment of F-35s in areas like Truax Field (Wisconsin), Hill Air Force Base (Utah), Burlington International Airport (Vermont), as well as some regions in Switzerland. These more recent noise “hotspots” have added to longstanding and persistent frustration with military aviation noise in Asia-Pacific regions (Baek et al., 2023; Jeong et al., 2012; Kim et al., 2014; Yokoshima et al., 2021), particularly the numerous airfields on the island of Okinawa (Tokuda and Barnett, 2017). Noise from military operations are not only a divisive public relations problem, but are likely to pose an increasing legal liability as urbanization continues to encroach on land near military bases (Appel and Smith, 2011; Kelly, 2019; Landis et al., 2001; Martens and Spyropoulos, 2010). We suggest that more proactive and adaptive approaches of noise management around airfields should include conducting regular noise monitoring and predicting population health outcomes of military aviation noise (Jacuzzi et al., 2024), as well as conducting surveys and analyzing complaint data to understand and address the priority and specific concerns in the community (Mabrey and Carey, 1980). Ideally, these activities would be conducted by federal agencies and airfields, but as our results demonstrate, communities can also effectively self-organize collection of complaint data to support advocacy.

## CRediT authorship contribution statement

**Ching-Hsuan Huang**: Conceptualization, Methodology, Software, Formal analysis, Investigation, Data curation, Writing – Original Draft. **Lauren M. Kuehne**: Conceptualization, Methodology, Software, Formal analysis, Investigation, Data curation, Writing – Original Draft, Project administration, Funding acquisition. **Giordano Jacuzzi:** Methodology, Writing – Reviewing and Editing. **Julian D. Olden**: Methodology, Writing – Reviewing and Editing, Funding acquisition, Resources. **Edmund Seto**: Conceptualization, Methodology, Writing – Reviewing and Editing, Funding acquisition, Resources, Supervision.

## Data Availability

All data produced in the present work are contained in the manuscript.

## Acknowledgements

We are grateful to Quiet Skies Over San Juan County and Citizens of Ebey’s Reserve for providing the noise complaint data for this study. Funding for this research was from the University of Washington Population Health Initiative. The study was evaluated using the self-determination process of the University of Washington Institutional Review Board (IRB) and was determined to be an exclusion that does not require IRB review.

## Supplementary Materials

**Figure S1.**
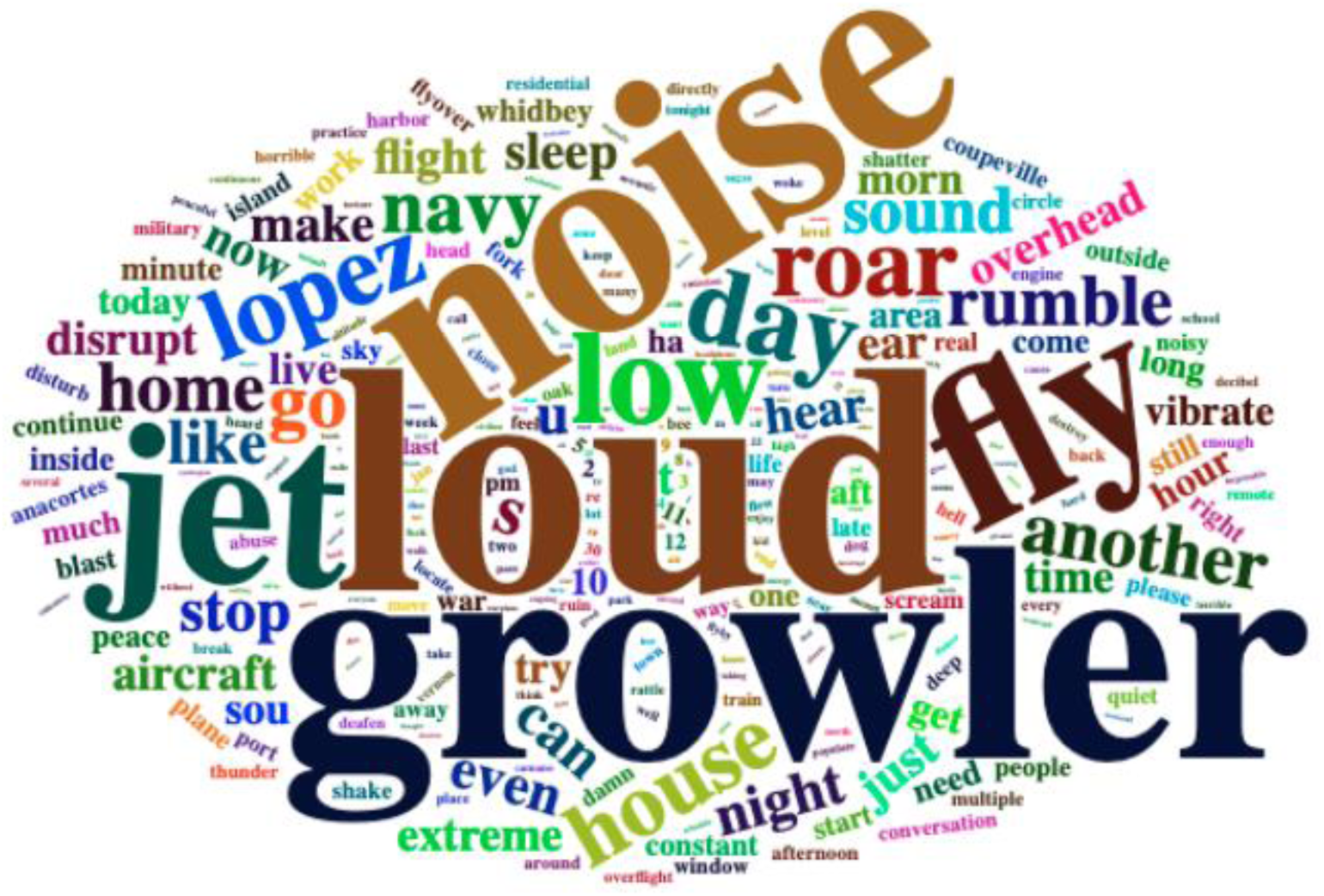
Word cloud of the high frequency words in the Quiet Skies complaints.

**Figure S2.**
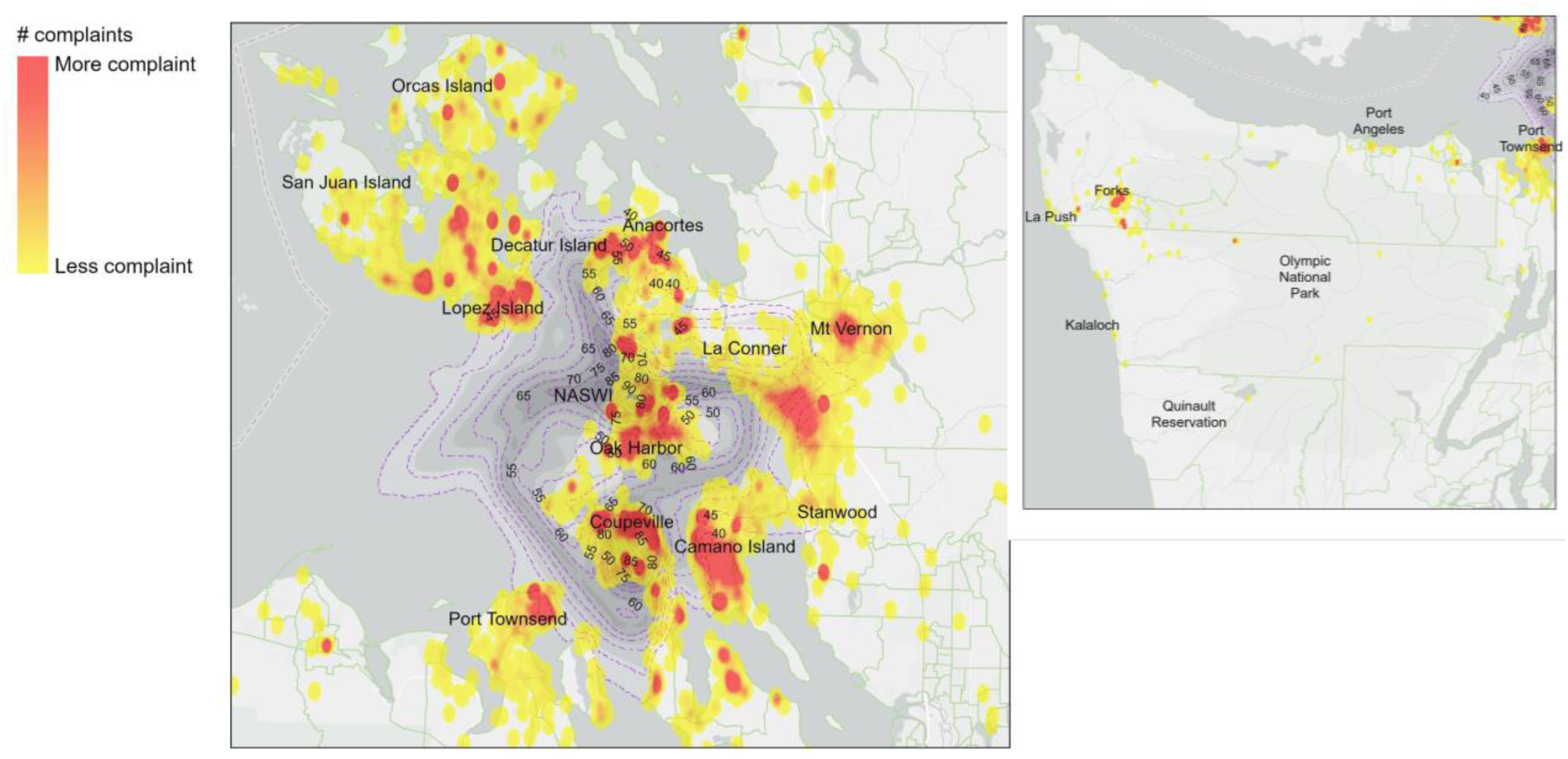
Kernel density of complaint quantity, overlaid with modeled day-night noise level (L_dn_ dBA). Left: NASWI Whidbey Island vicinity. Right: Olympic Peninsula.

**Figure S3.**
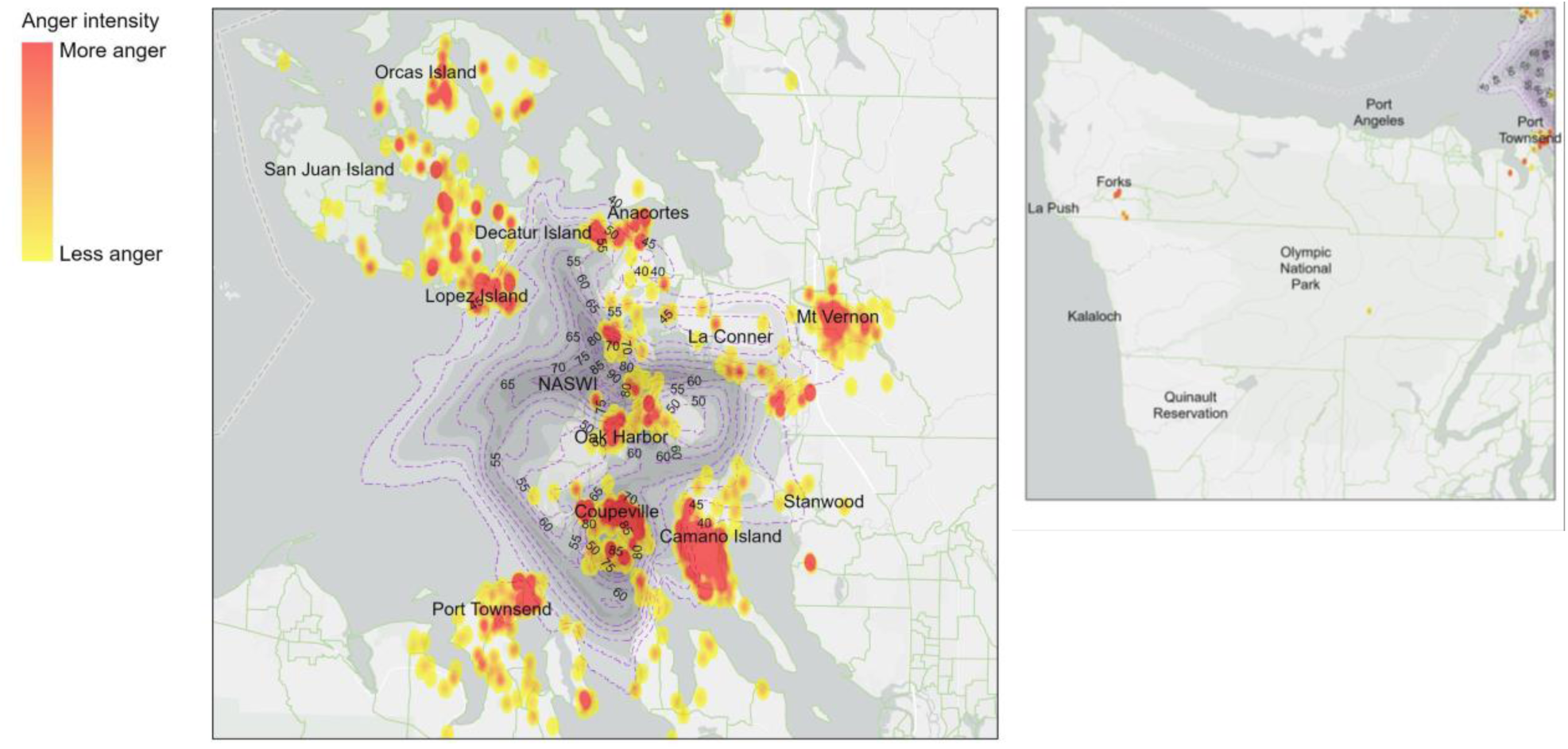
Kernel density of anger emotion intensity in complaints, overlaid with modeled day-night noise level (L_dn_ dBA). Left: NASWI Whidbey Island vicinity. Right: Olympic Peninsula.

**Figure S4.**
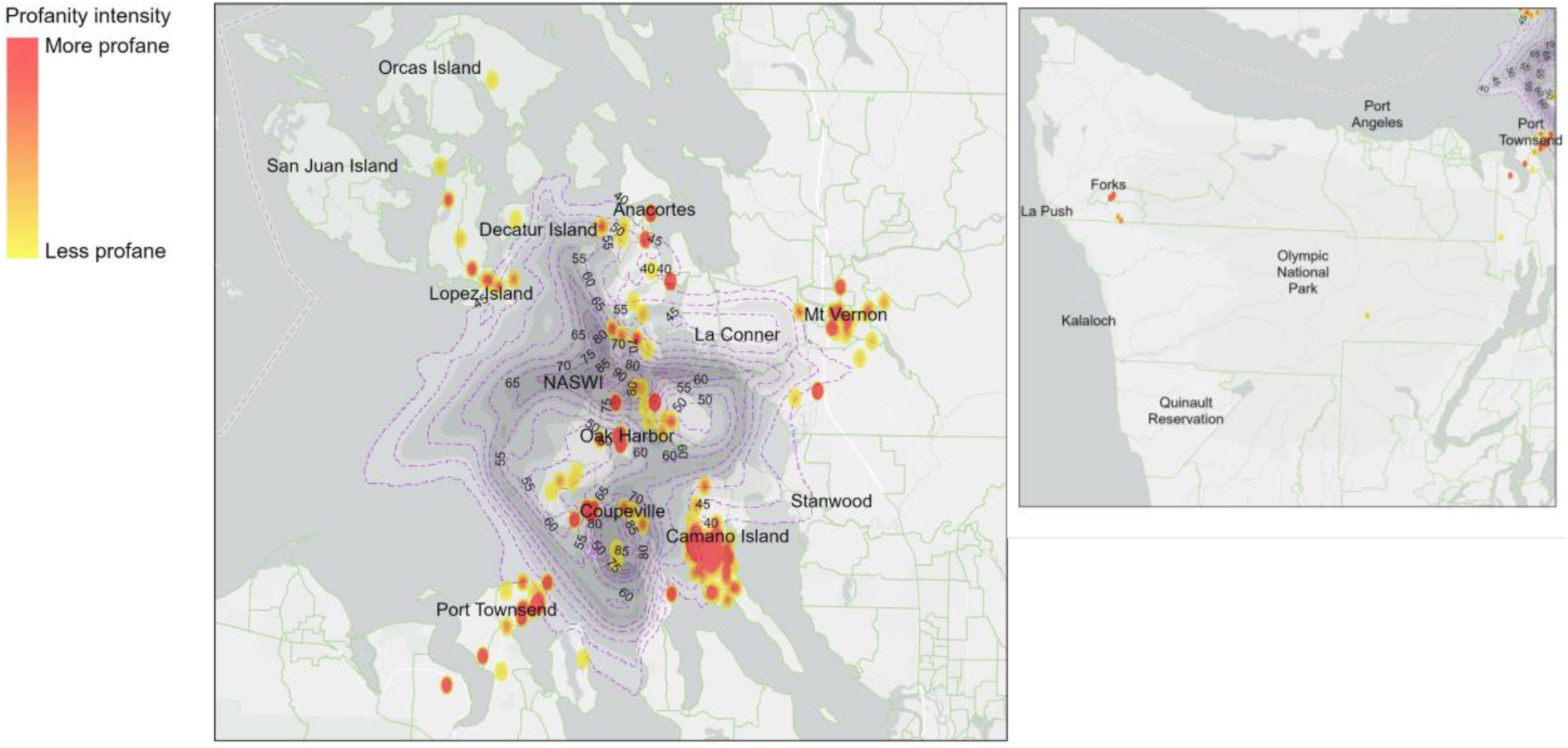
Kernel density of profane emotion intensity in complaints, overlaid with modeled day-night noise level (L_dn_ dBA). Left: NASWI Whidbey Island vicinity. Right: Olympic Peninsula.

**Figure S5.**
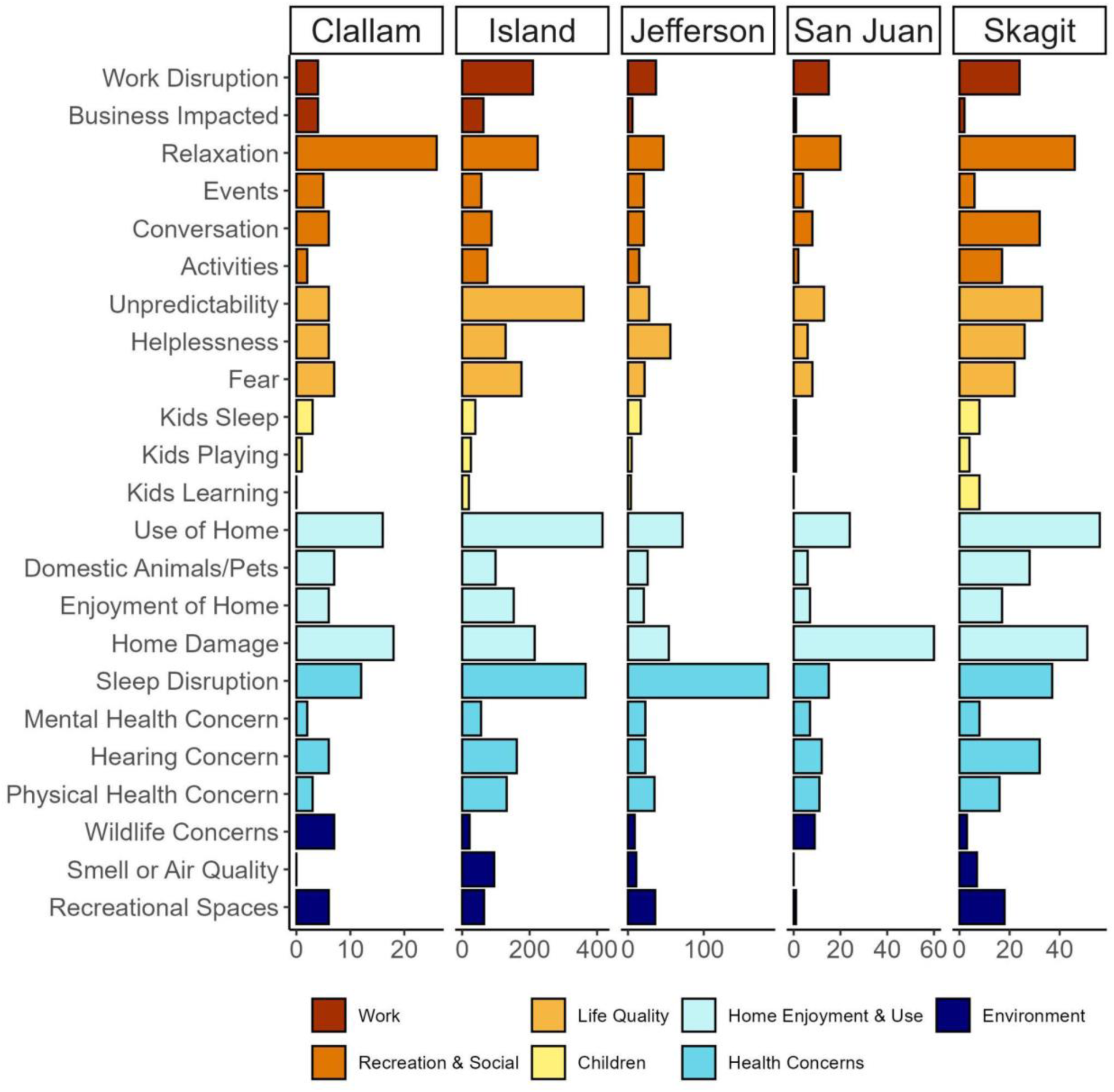
Complaint classifications for Navy line complaints, for the five most impacted counties (*n*=1,978). Complaints were classified into 23 subcategories within seven broad categories (colored bars) describing impacts on health and quality of life.

**Figure S6.**
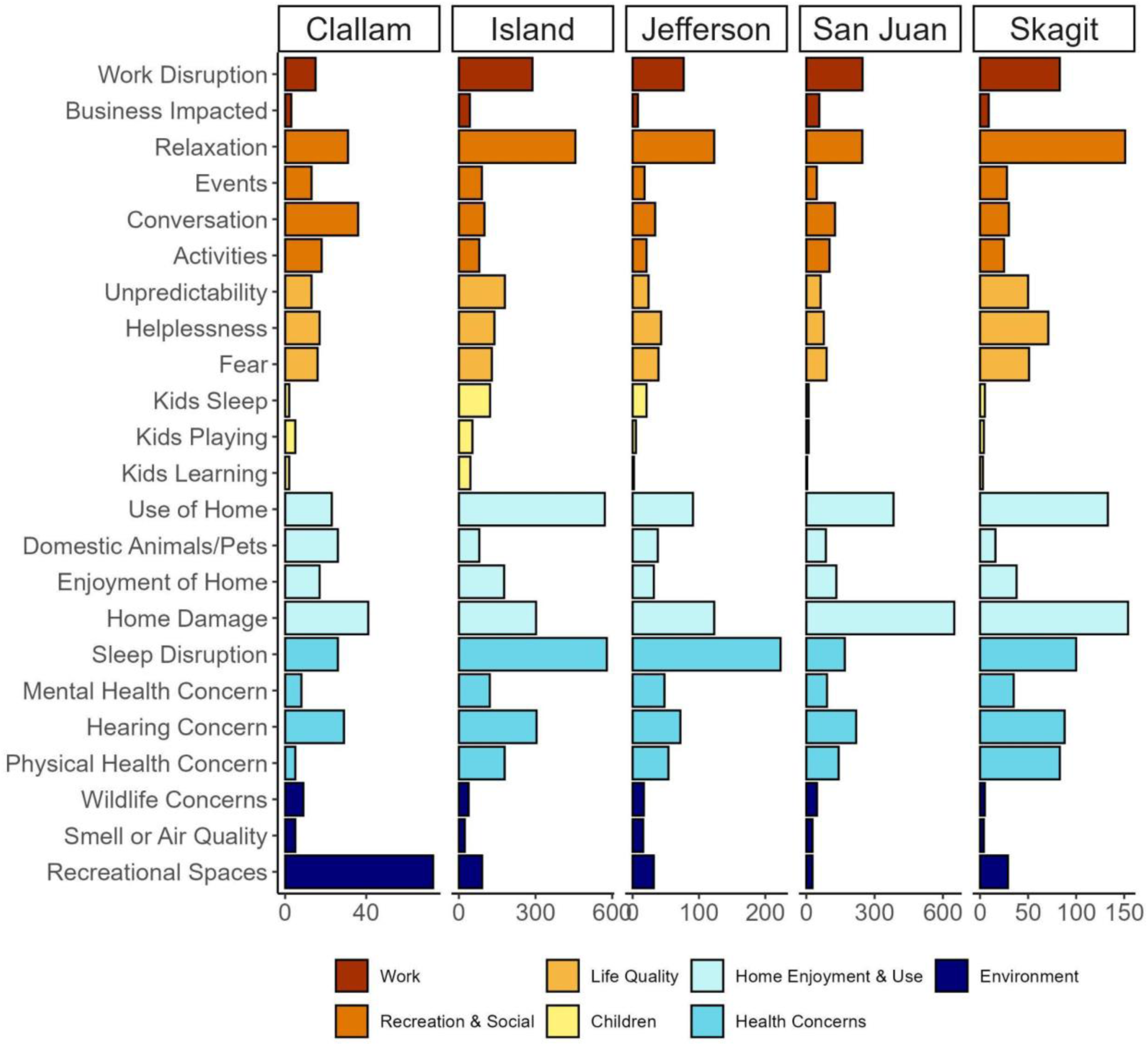
Complaint classifications for Quiet Skies complaints, for the five most impacted counties (*n*=3,933). Complaints were classified into 23 subcategories within seven broad categories (colored bars) describing impacts on health and quality of life.

**Table S1.**
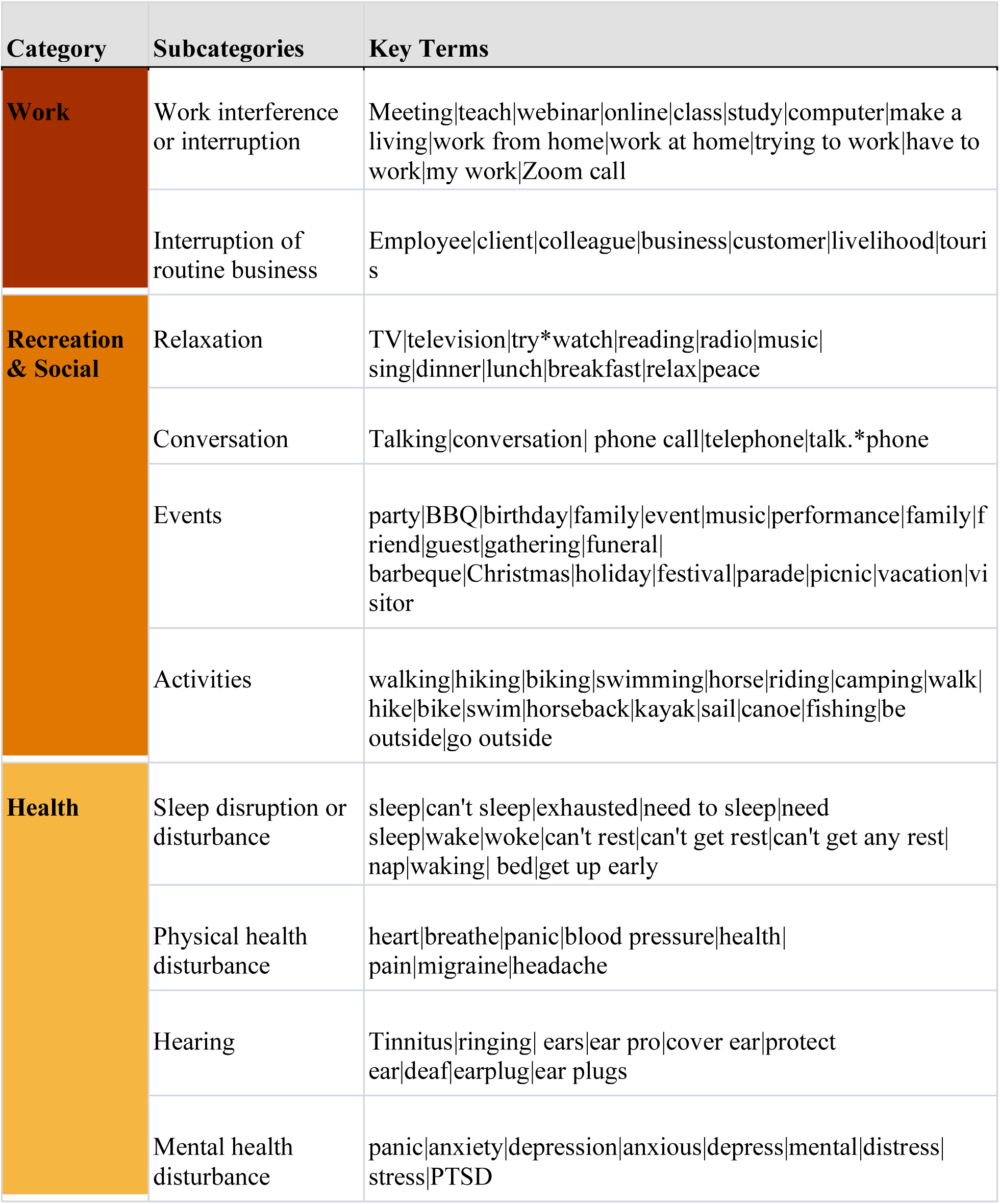
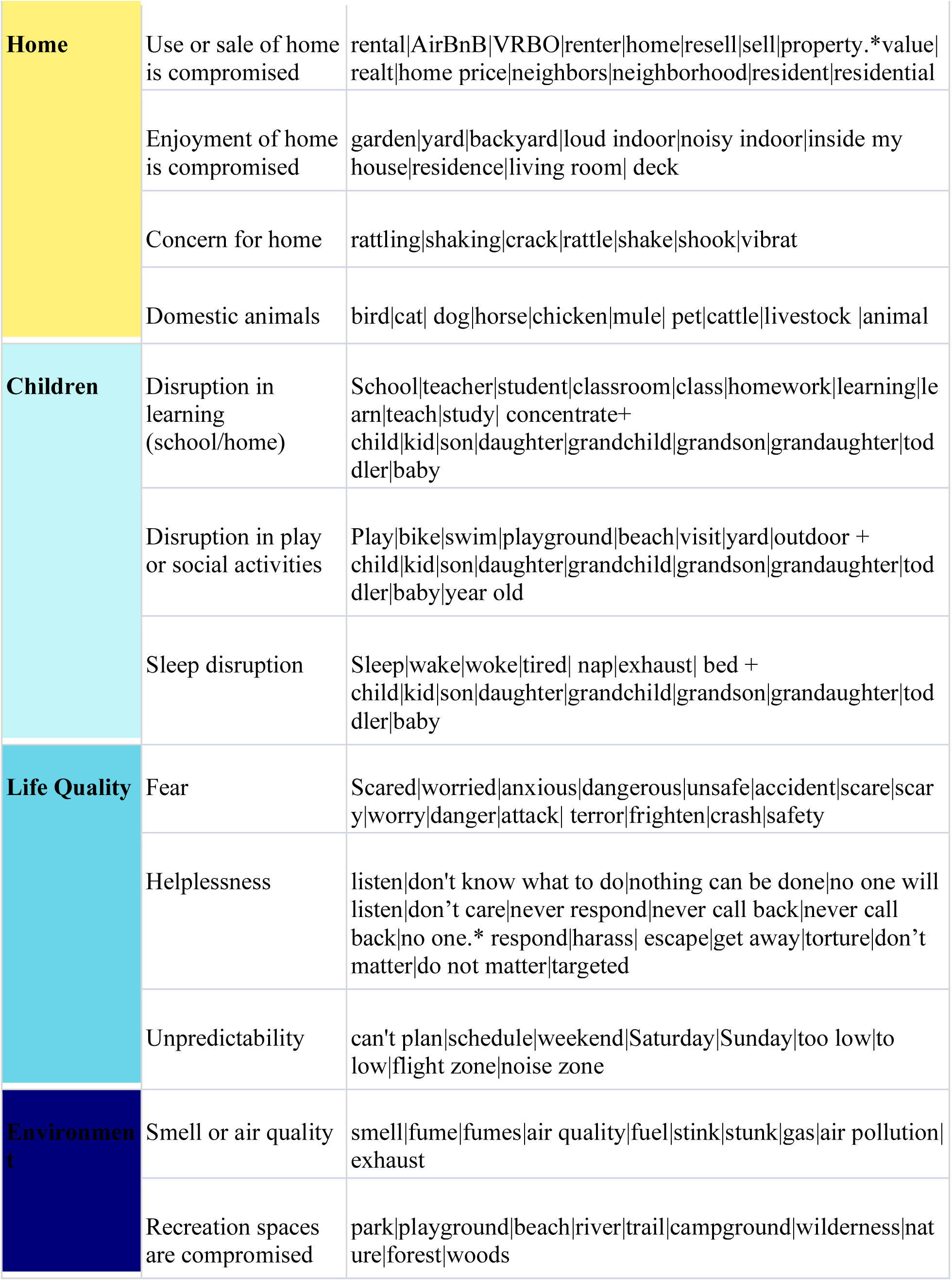
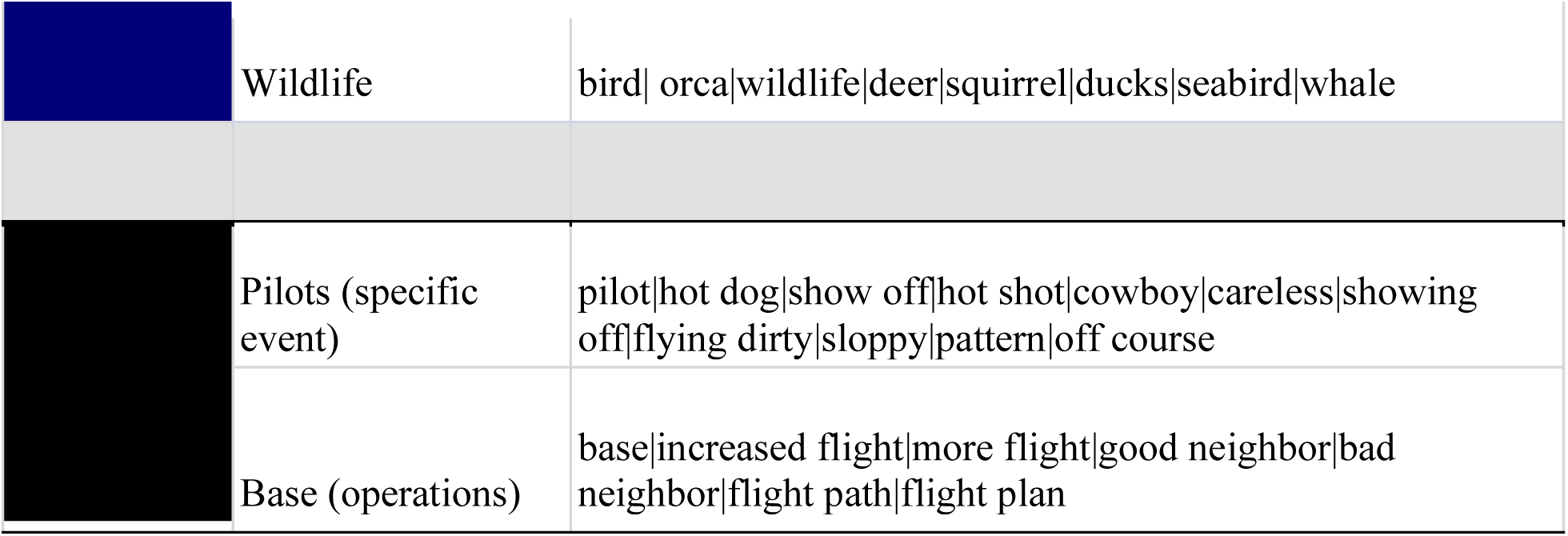
Complaint taxonomy and key term approach of 23 subcategories (within seven categories) to classify complaints (0=subcategory is not represented, 1=subcategory is represented) based on Key Terms within the comment. Some terms were better captured by stems than complete words (e.g., “vibrat” vs. “vibration”). A “+” indicates a combined term, and a “.*” indicates a wildcard joining two terms with indeterminate words between.

**Table S2.**
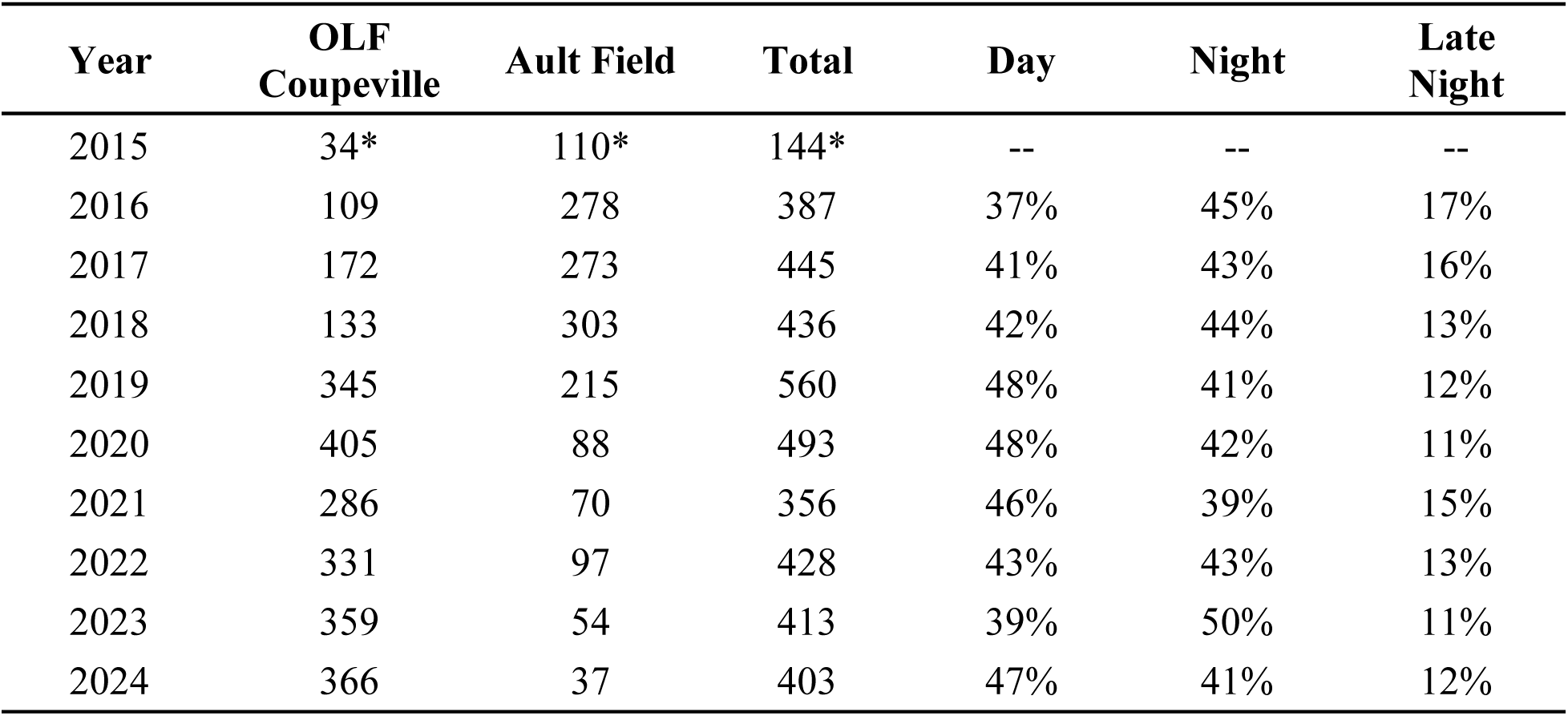
Annual summary of scheduled field carrier landing practice (FCLP) for Ault Field and Coupeville OLF for the years 2015-2023. All years except 2015 were based on FCLP schedule notifications, which are published by NASWI for the coming week. Following the protocols established in Kuehne and Olden (2020), we summarized the weekly notification schedule for FCLPs out of Naval Air Station Whidbey Island to establish patterns over the past nine years (U.S. Dept of the Navy 2019). FCLP notifications are published using consistent descriptions that allow classification as occurring in one of nine timeframes: Early AM, Late AM, Early PM, Late PM, Early Evening, Evening, Night, Late Night, Past Midnight. The percentage of FCLP sessions for the three periods used in modeling complaints are shown: Day (timeframes = Early/Late Morning, Early/Late Afternoon), Night (timeframes = Early/Late Evening, Night), and Late Night (timeframes = Late Night, Past Midnight).

| **Year** | **OLF**<br>**Coupeville** | **Ault Field** | **Total** | **Day** | **Night** | **Late**<br>**Night** |
| --- | --- | --- | --- | --- | --- | --- |
| 2015 | 34* | 110* | 144* | -- | -- | -- |
| 2016 | 109 | 278 | 387 | 37% | 45% | 17% |
| 2017 | 172 | 273 | 445 | 41% | 43% | 16% |
| 2018 | 133 | 303 | 436 | 42% | 44% | 13% |
| 2019 | 345 | 215 | 560 | 48% | 41% | 12% |
| 2020 | 405 | 88 | 493 | 48% | 42% | 11% |
| 2021 | 286 | 70 | 356 | 46% | 39% | 15% |
| 2022 | 331 | 97 | 428 | 43% | 43% | 13% |
| 2023 | 359 | 54 | 413 | 39% | 50% | 11% |
| 2024 | 366 | 37 | 403 | 47% | 41% | 12% |

## References

1. Appel, R., Smith, T., 2011. Compatible Land Use Near Military Installations: The Problem and the Local Government Response. Planning & Environmental Law 63, 3–10. 10.1080/15480755.2011.569207

2. Baek, K., Park, C., Sakong, J., 2023. The Impact of Aircraft Noise on the Cognitive Function of Elementary School Students in Korea. Noise and Health 25, 83–91. 10.4103/nah.nah_71_22

3. Banks, J.L., O’Rourke, B.P., 2025. Community impacts of aviation noise: a pilot survey. J Expo Sci Environ Epidemiol. 10.1038/s41370-024-00737-z

4. Basner, M., Babisch, W., Davis, A., Brink, M., Clark, C., Janssen, S., Stansfeld, S., 2014. Auditory and non-auditory effects of noise on health. The Lancet 383, 1325–1332.

5. Bell, W.Y., 1956. A study of community reactions to the Weymouth Naval Air Station and its jet aircraft operations. (PhD Thesis). Boston University.

6. Beranek, L., 1952. Jet Airports May Be 100 Times Noisier. Science Digest XXXII, 24.

7. Borsky, P., 1961a. Community Reactions to Air Force noise. Part I. Basic concepts and preliminary methodology. National Opinion Research Center, University of Chicago, Chicago, IL.

8. Borsky, P., 1961b. Community Reactions to Air Force noise. Part II. Data on community studies and their interpretation. National Opinion Research Center, University of Chicago, Chicago, IL.

9. Bouchet-Valat, M., Porter, M., Boulton, R., 2023. SnowballC: Snowball Stemmers Based on the C “libstemmer” UTF-8 Library.

10. Bowen, D., Geng, L., 2017. Having a Lot Isn’t Enough: Trends in Upsizing Houses and Shrinking Lots. FEDS Notes. Washington: Board of Governors of the Federal Reserve System.

11. Buxton, R.T., McKenna, M.F., Mennitt, D., Brown, E., Fristrup, K., Crooks, K.R., Angeloni, L.M., Wittemyer, G., 2019. Anthropogenic noise in US national parks–sources and spatial extent. Front Ecol Environ 17, 559–564.

12. Clark, C., Gjestland, T., Lavia, L., Notley, H., Michaud, D., Morinaga, M., 2021. Assessing community noise annoyance: A review of two decades of the international technical specification ISO/TS 15666: 2003. The Journal of the Acoustical Society of America 150, 3362–3373.

13. Dreier, C., Vorländer, M., 2021. Aircraft noise—Auralization-based assessment of weather-dependent effects on loudness and sharpness. The Journal of the Acoustical Society of America 149, 3565–3575. 10.1121/10.0005040

14. Efroymson, R.A., Suter II, G.W., Rose, W.H., Nemeth, S., 2001. Ecological risk assessment framework for low-altitude aircraft overflights: I. Planning the analysis and estimating exposure. Risk Analysis 21, 251–262.

15. El Barachi, M., AlKhatib, M., Mathew, S., Oroumchian, F., 2021. A novel sentiment analysis framework for monitoring the evolving public opinion in real-time: Case study on climate change. Journal of Cleaner Production 312, 127820. 10.1016/j.jclepro.2021.127820

16. Fan, Y., Teo, H.P., Wan, W.X., 2021. Public transport, noise complaints, and housing: Evidence from sentiment analysis in Singapore. Journal of Regional Science 61, 570–596. 10.1111/jors.12524

17. Fidell, S., 2003. The Schultz curve 25 years later: A research perspective. The Journal of the Acoustical Society of America 114, 3007–3015. 10.1121/1.1628246

18. Fidell, S., Howe, R., 1998. Use of airport noise complaint files to improve understanding of community response to aircraft noise.

19. Fidell, S., Mestre, V., Sneddon, M., 2012. A potential role for noise complaints as a predictor of the prevalence of annoyance with aircraft noise. noise cont engng j 60, 62–68. 10.3397/1.3677792

20. Gasco, L., Schifanella, R., Aiello, L.M., Quercia, D., Asensio, C., De Arcas, G., 2020. Social Media and Open Data to Quantify the Effects of Noise on Health. Front. Sustain. Cities 2, 41. 10.3389/frsc.2020.00041

21. Gelderblom, F.B., Gjestland, T.T., Granoien, I.L., Taraldsen, G., 2014. The impact of civil versus military aircraft noise on noise annoyance, in: INTER-NOISE and NOISE-CON Congress and Conference Proceedings. Institute of Noise Control Engineering, pp. 786–795.

22. Guski, R., Felscher-Suhr, U., Schuemer, R., 1999. The concept of noise annoyance: how international experts see it. Journal of Sound and Vibration 223, 513–527. 10.1006/jsvi.1998.2173

23. Hadley, W., 2023. stringr: Simple, Consistent Wrappers for Common String Operations.

24. Hahsler, M., Piekenbrock, M., Arya, S., Mount, D., Malzer, C., 2023. dbscan: Density-Based Spatial Clustering of Applications with Noise (DBSCAN) and Related Algorithms.

25. Hart, C.W., Goldstein, H., Sheatsley, P.B., 1954. Community aspects of aircraft annoyance (No. 54). National Opinion Research Center, Chicago, IL.

26. Hu, M.-C., Pavlicova, M., Nunes, E.V., 2011. Zero-Inflated and Hurdle Models of Count Data with Extra Zeros: Examples from an HIV-Risk Reduction Intervention Trial. The American Journal of Drug and Alcohol Abuse 37, 367–375. 10.3109/00952990.2011.597280

27. Hume, K., Gregg, M., Thomas, C., Terranova, D., 2003. Complaints caused by aircraft operations: an assessment of annoyance by noise level and time of day. Journal of Air Transport Management 9, 153–160. 10.1016/S0969-6997(02)00079-0

28. Hume, K., Terranova, D., Thomas, C., 2002. Complaints and annoyance caused by aircraft operations: Temporal patterns and individual bias. Noise and Health 4, 45–55.

29. Ising, H., Rebentisch, E., Babisch, W., Curio, I., Sharp, D., Baumgärtner, H., 1990. Medically relevant effects of noise from military low-altitude flights—results of an interdisciplinary pilot study. Environment International 16, 411–423.

30. Jacuzzi, G., Kuehne, L.M., Harvey, A., Hurley, C., Wilbur, R., Seto, E., Olden, J.D., 2024. Population health implications of exposure to pervasive military aircraft noise pollution. J Expo Sci Environ Epidemiol. 10.1038/s41370-024-00670-1

31. Jeong, Y.-R., Park, J.-B., Min, K.-B., Lee, C., Kil, H.-G., Lee, W.-W., Lee, K.-J., 2012. The effects of aircraft noise exposure upon hearing loss, anxiety, and depression on subjects residing adjacent to a military airbase. Korean Journal of Occupational and Environmental Medicine 24, 40–51.

32. Kelly, J.K., 2019. The Sound of Freedom at Naval Air Station Whidbey: Environmental Impact Review under the National Historic Preservation Act and National Environmental Policy Act. Villanova Environmental Law Journal 31, 113.

33. Kerry, G., Weeler, P.D., Hempstock, T.I., James, D.J., 1998. Impulse noise metrics and their application to noise from low flying military jet aircraft. The Journal of the Acoustical Society of America 103, 2800–2800.

34. Kim, S.J., Chai, S.K., Lee, K.W., Park, J.-B., Min, K.-B., Kil, H.G., Lee, C., Lee, K.J., 2014. Exposure–response relationship between aircraft noise and sleep quality: A community-based cross-sectional study. Osong public health and research perspectives 5, 108–114.

35. Kuehne, L.M., Erbe, C., Ashe, E., Bogaard, L.T., Salerno Collins, M., Williams, R., 2020. Above and below: military aircraft noise in air and under water at Whidbey Island, Washington. Journal of Marine Science and Engineering 8, 923.

36. Kuehne, L.M., Hurley, C., In Press. Military noise as a health threat, in: Birch, M., Hagopian, A. (Eds.), Handbook of Conflict and Health, De Gruyter Contemporary Social Sciences Handbooks. De Gruyter, Berlin, Germany.

37. Kuehne, L.M., Olden, J.D., 2020. Military flights threaten the wilderness soundscapes of the Olympic Peninsula, Washington. Northwest Science 94, 188–202.

38. Landis, J.D., Foster, H., Frontiera, P., Reilly, M., Twiss, R., 2001.Forecasting and Mitigating Future Urban Encroachment Adjacent to California Military Installations: A Spatial Approach.

39. Mabrey, J.L., Carey, R.B., 1980. An analysis of community complaints to Air Force aircraft noise. Air Force Aerospace Medical Research Laboratory, Wright Patterson Air Force Base, Ohio.

40. Mabry, J.E., Carey, R.B., 1980. An Analysis of Community Complaints to Air Force Aircraft Noise.

41. Mace, B.L., Bell, P.A., Loomis, R.J., 1999. Aesthetic, Affective, and Cognitive Effects of Noise on Natural Landscape Assessment. Society & Natural Resources 12, 225–242. 10.1080/089419299279713

42. Mahl, D., Guenther, L., 2023. Content Analysis in the Research Field of Environmental & Climate Change Coverage, in: Oehmer-Pedrazzi, F., Kessler, S.H., Humprecht, E., Sommer, K., Castro, L. (Eds.), Standardisierte Inhaltsanalyse in der Kommunikationswissenschaft – Standardized Content Analysis in Communication Research. Springer Fachmedien Wiesbaden, Wiesbaden, pp. 203–212. 10.1007/978-3-658-36179-2_18

43. Martens, S., Spyropoulos, J.T., 2010. Practical jet noise reduction for tactical aircraft, in: Turbo Expo: Power for Land, Sea, and Air. pp. 389–399.

44. Montini, T., Noble, A.A., Stelfox, H.T., 2008. Content analysis of patient complaints. International Journal for Quality in Health Care 20, 412–420. 10.1093/intqhc/mzn041

45. Okinawa Prefectural Government, 1999. A Report on the Aircraft Noise as a Public Health Problem in Okinawa. Department of Culture and Environmental Affairs, Okinawa, Japan.

46. Olden, J.D., Kuehne, L.M., Kim, A., Jacuzzi, G., Seto, E., In Review. Noise-induced annoyance and sleep disturbance from military aircraft training. BMC Public Health.

47. Pepper, C.B., Nascarella, M.A., Kendall, R.J., 2003. A review of the effects of aircraft noise on wildlife and humans, current control mechanisms, and the need for further study. Environmental Management 32, 418–432.

48. Pinheiro, J.C., Bates, D.M., 2000. Mixed-effects models in S and S-PLUS, Statistics and computing. Springer, New York.

49. R Core Team, 2025.R: A language and environment for statistical computing.

50. Reader, T.W., Gillespie, A., Roberts, J., 2014. Patient complaints in healthcare systems: a systematic review and coding taxonomy. BMJ quality & safety 23, 678–689.

51. Rinker, T.W., 2021a. sentimentr: Calculate Text Polarity Sentiment. Buffalo, New York.

52. Rinker, T.W., 2021b. lexicon: Lexicon Data.

53. Silge, J., Robinson, D., 2016. tidytext: Text Mining and Analysis Using Tidy Data Principles in R. JOSS 1(3). doi:10.21105/joss.00037

54. Stansfeld, S.A., Matheson, M.P., 2003. Noise pollution: non-auditory effects on health. British Medical Bulletin 68, 243–257. 10.1093/bmb/ldg033

55. Stusnick, E., Bradley, K.A., Bossi, M.A., Molino, J.A., Rickert, D.G., 1993. The Effect of Onset Rate on Aircraft Noise Annoyance. Volume 3. Hybrid Own-Home Experiment (Final Report No. AFRL-HE-WP-TR-2001-0130). United States Air Force Research Laboratory, Alexandria, VA.

56. Tokuda, Y., Barnett, P.B., 2017. Constructing a new US Military Base: a health threat to Okinawan people. Environmental Justice 10, 23–25.

57. Tsukioka, H., Morinaga, M., Yamamoto, I., Yamada, I., Hinai, T., 2011. Review of environmental measures for noise issues around defense facilities in Japan, in: INTER-NOISE and NOISE-CON Congress and Conference Proceedings. Institute of Noise Control Engineering, pp. 3409–3416.

58. US Dept of the Navy, 2025a. Final Amended Analysis to the Environmental Impact Statement for EA-18G “Growler” Airfield Operations at Naval Air Station Whidbey Island Complex, Washington, September 2018. United States Department of the Navy, Washington D.C.

59. US Dept of the Navy, 2025b. Naval Air Station Whidbey Island (NASWI) Noise Complaint System.

60. US Dept of the Navy, 2020. Northwest Training and Testing Final Supplemental Environmental Impact Statement/Overseas Environmental Impact Statement (FEIS/OEIS). United States Department of the Navy, Washington D.C.

61. US Environmental Protection Agency, 1971. The Social Impact of Noise (No. NTID300.11). US Environmental Protection Agency, Washington D.C.

62. van Wiechen, C.M.A.G., Franssen, E.A.M., de Jong, R.G., Lebret, E., 2002. Aircraft Noise Exposure from Schiphol Airport : A Relation with Complainants. Noise and Health 5.

63. Waitz, I.A., Lukachko, S.P., Lee, J.J., 2005. Military aviation and the environment: Historical trends and comparison to civil aviation. Journal of Aircraft 42, 329–339.

64. Wang, X., Zhu, Y., Zeng, H., Cheng, Q., Zhao, X., Xu, H., Zhou, T., 2022. Spatialized Analysis of Air Pollution Complaints in Beijing Using the BERT+CRF Model. Atmosphere 13, 1023. 10.3390/atmos13071023

65. Weinandy, R., Myck, T., Thierbach, R., 2014. Land-use planning at airports in Germany, in: INTER-NOISE and NOISE-CON Congress and Conference Proceedings. Institute of Noise Control Engineering, pp. 157–161.

66. Wemheuer, R.F., 1979. Analysis of the Relationship Between Community Attitude Toward Military Aircraft Noise, Safety and Economic Factors Associated with the Operation of Marine Corps Air Station, El Toro, California, and the Community Attitude Toward the Continued Operation of the Installation. Pepperdine University.

67. Wickham, H., 2016. ggplot2: Elegant Graphics for Data Analysis.

68. World Health Organization, 2022. Compendium of WHO and other UN guidance on health and environment. Chapter 11. Environmental noise (No. WHO/HEP/ECH/EHD/22.01). World Health Organization, Geneva, Switzerland.

69. World Health Organization, 2011. Burden of disease from environmental noise: Quantification of healthy life years lost in Europe. World Health Organization. Regional Office for Europe, Copenhagen, Denmark.

70. Yokoshima, S., Morinaga, M., Tsujimura, S., Shimoyama, K., Morihara, T., 2021. Representative Exposure–Annoyance Relationships Due to Transportation Noises in Japan. International journal of environmental research and public health 18, 10935.

71. Zambon, G., Muchetti, S.S., Salvi, D., Angelini, F., Brambilla, G., Benocci, R., 2020. Analysis of noise annoyance complaints in the city of Milan, Italy. J. Phys.: Conf. Ser. 1603, 012029. 10.1088/1742-6596/1603/1/012029

